# Automated Classification of At-home SARS-CoV-2 Lateral Flow Assay Test Results using Image Matching and Transfer Learning: multiple-pipeline study

**DOI:** 10.1101/2024.01.04.24300836

**Authors:** Meysam Safarzadeh, Carly Herbert, Steven Koon Wong, Pamela Stamegna, Yurima Guilarte-Walker, Colton Wright, Thejas Suvarna, Chris Nowak, Vik Kheterpal, Shishir Pandey, Biqi Wang, Honghuang Lin, Laurel O’Connor, Nathaniel Hafer, Katherine Luzuriaga, Yuka Manabe, John Broach, Adrian H Zai, David D McManus, Xian Du, Apurv Soni

## Abstract

**Introduction:** Rapid antigen testing for SARS-CoV-2 is an important tool for the timely diagnosis of COVID-19, especially in at-home settings. However, the interpretation of test results can be subjective and prone to error. We describe an automated image analysis pipeline to accurately classify test types and results without human intervention using a dataset of 51,274 rapid antigen test images across three distinct test card brands.

**Methods:** The proposed method classifies participant-submitted images into four categories: positive for SARS-CoV-2, negative for SARS-CoV-2, invalid/uncertain, and unclassifiable. The model includes four stages: test card classification and region of interest detection using image-matching algorithms, elimination of invalid results using a developed Siamese neural network, and test result classification using transfer learning.

**Results:** The model accuracy was very good for test-card classification (100%), region of interest detection (83.5%), and identification of invalid results ranging from 95.6% to 100% for different test types. Performance of the model for test result classification varied by tests; the model’s sensitivity, specificity, and precision for Abbott BinaxNOW™ was 0.761, 0.989, and 0.946, BD Veritor™ At-Home COVID-19 Test was 0.955, 0.993, and 0.877, and for QuickVue® At-Home OTC COVID-19 Test was 0.816, 0.988, and 0.930.

**Conclusion:** The proposed method improved the interpretation of rapid antigen tests, particularly in invalid result detection compared to human-read, and offers a great opportunity for standardization of rapid antigen test interpretation and for providing feedback to participants with invalid tests.

## INTRODUCTION

The COVID-19 pandemic has brought about an urgent need for the development of effective diagnostic tools to control the rapid spread of the virus through early detection and isolation [1] [2]. In response, the Food and Drug Administration has issued Emergency Use Authorizations (EUAs) for many Antigen-detection rapid diagnostic tests (Ag-RDTs) for COVID-19 [3], [4]. These tests are approved for home use and yield results within 15 minutes, making them easy to use, fast, and widely accessible. However, they have been found to have a higher rate of false negatives compared to molecular tests [5]. Ensuring the proper interpretation of Ag-RDT results is therefore critical to preserve their sensitivity to the greatest possible extent.

Interpretation issues, such as weak or faint test lines, and bias due to the presence or absence of symptoms or close contacts, can impede the accurate interpretation of results. Furthermore, invalid results, which occur when the test card displays anomalous line shapes, can potentially be misinterpreted as positive or negative results by patients. Additionally, people with varying degrees of vision impairment may face difficulty interpreting the results [6]. Thus, efforts to minimize false negative and false positive rates are essential. Artificial intelligence (AI) has been proposed as a promising solution to reduce interpretation issues associated with at-home diagnostics [7] [8] [9]. However, prior studies and algorithms are typically test-specific and were performed in controlled laboratory settings, which imposes additional constraints on end-users to identify proper lighting and background when capturing images. In contrast, this study examined the effect of different test cards on the model’s performance, detection and prevalence of invalid test results, and ways to improve the readability of test cards obtained from users in non-lab settings.

In this paper, we present an automatic and intelligent framework that utilizes advanced machine learning techniques and image-matching algorithms to interpret at-home SARS-Cov-2 Ag-RDTs. Our proposed approach is based on a dataset of 51,274 images of test cards submitted by participants as part of the Rapid Acceleration of Diagnostics (RADx®) projects. The proposed method consists of four consecutive modules: 1) test card classification; 2) detection and cropping of the region of interest (ROI); 3) invalid result detection; 4) ROI classification module to classify ROIs into negative and positive results with corresponding confidence.

## MATERIAL AND METHODS

### Dataset

This model was built and tested using rapid antigen test images from participants enrolled in digital cohort studies funded by the Rapid Acceleration of Diagnostics (RADx) program of the National Institutes of Health. Participants over 2 years of age living in the mainland United States with access to a smartphone were eligible to participate. More details about these studies and the complete inclusion criteria can be found elsewhere [10], [11]. Participants provided written informed consent to participate in the study through a mobile app, and all study procedures were conducted using the app. Participants were assigned to receive rapid antigen tests from one of the three brands (BinaxNOW™, BD Veritor™, and QuickVue®) shown in Fig. S1. Participants received shipments of rapid antigen tests to their homes and were instructed to complete periodic rapid antigen tests throughout the study period. During each testing period, the participants were instructed to use the tests according to the instructions indicated per emergency use authorization. Participants were asked to indicate the results of their test (positive, negative, invalid, or do not know) in the app and upload a photo of their test strip to the app. The study coordinators read and interpreted each test using the uploaded images to provide expert reads. The RADx studies and the use of these images were approved by Western Copernicus Group (WCG) IRB.

### Test Card Classification and ROI Detection

The proposed card interpretation system follows a sequential process, outlined in Fig. 1. Firstly, an image matching algorithm is employed to determine the type of test card, such as Quidel QuickVue®, Abbott BinaxNOW™, or BD Veritor™. Secondly, the region of interest (ROI) on the test card is detected and cropped using the homography matrix. An example of the ROI, specifically the result window, is illustrated in Fig. 1. Also, Fig. S2 provides an in-depth depiction of the workflow covering the Test card classification and ROI detection stages. To classify the test cards, advanced image-matching techniques were utilized, which offer valuable transformation information for the detected objects, thereby facilitating subsequent ROI detection and cropping. We conducted evaluations using three state-of-the-art feature matching techniques, namely SIFT [12], SuperPoint + SuperGlue [13], [14], and ORB + BEBLID [15], [16] with various configurations. Following the feature matching process, we assessed the performance of outlier feature removal algorithms including MAGSAC++, GCRANSAC [26], and DEGENSAC [11]. In the third step of the workflow, the Invalid Result Detection module plays a crucial role in identifying and removing invalid ROIs. To accomplish this, two algorithms, namely the Siamese network and Big Transfer, were developed and rigorously evaluated. Following the elimination of invalid ROIs, the remaining valid ROIs undergo classification as whether positive, negative, or uncertain results. This classification task is carried out using convolutional neural network (CNN)-based models, specifically ResNet 50v2 and Big Transfer. Furthermore, the classification process includes the computation of a confidence score or measure associated with each classified ROI.

**Fig. 1.**
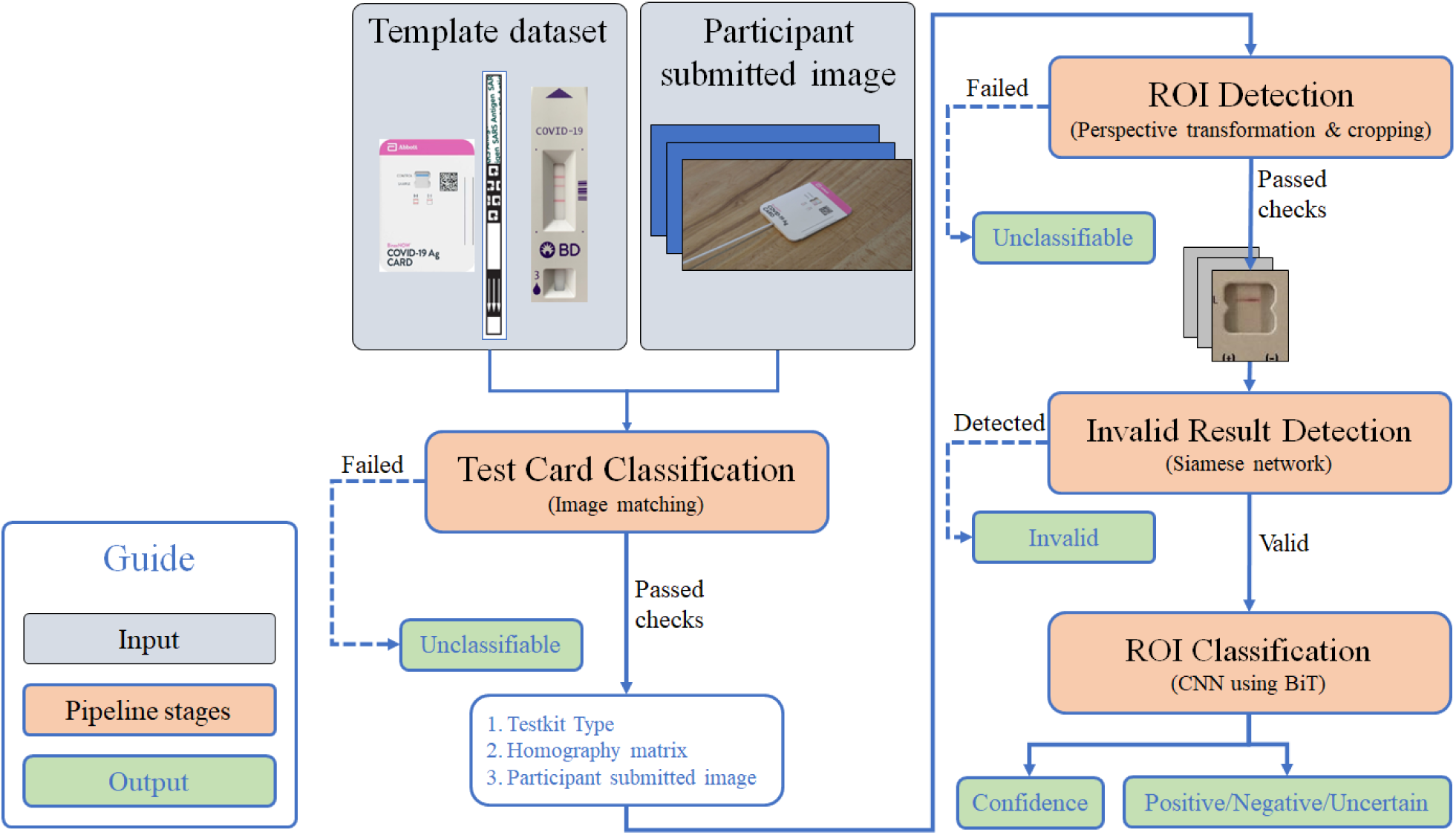
The workflow of the proposed method.

### Invalid Result Detection

We designed a Siamese network to distinguish between valid and invalid samples. Invalid ROIs included unused or faulty tests that lacked a control line (shown in Fig. S3). This model, illustrated in Fig. S4 which uses m-r50×1 backbone [17] as a feature extractor, facilitates the calculation of similarity between the input ROI and each invalid template image (see Fig. S5). If the similarity exceeds a certain threshold, the input sample was classified as invalid and discarded. More detailed explanations and configerations are available in Supplementary Methods and Table S1.

### ROI Classification

The ROI Classification module was developed with the objective of classifying the test result as either negative or positive, along with providing a confidence score for its prediction. We utilized transfer learning, as many well-trained models that already have shown their capability in object classification [17]. Specifically, we finetuned ResNet 50V2 [18] and Big transfer [17] (m-r50×1) pretrained on ImageNet. Further information is available in Supplementry Methods.

## RESULTS

### Sample size and Data Characteristics

The dataset distribution flowchart is depicted in Fig. 2. In this study, we employed a dataset comprising 51,274 images of rapid antigen tests collected from 6,952 participants. The dataset encompassed images obtained from three different test brands, namely Quidel QuickVue®, BD Veritor™, and Abbott BinaxNOW™. Within our collection of images, a total of 877 samples, equivalent to 1.7% of the entire dataset, were identified as positive for SARS-CoV-2 through the assessment of expert clinicians involved in the study. Images lacking a test card were excluded from the dataset for subsequent analyses, resulting in Subset 1.

**Fig. 2.**
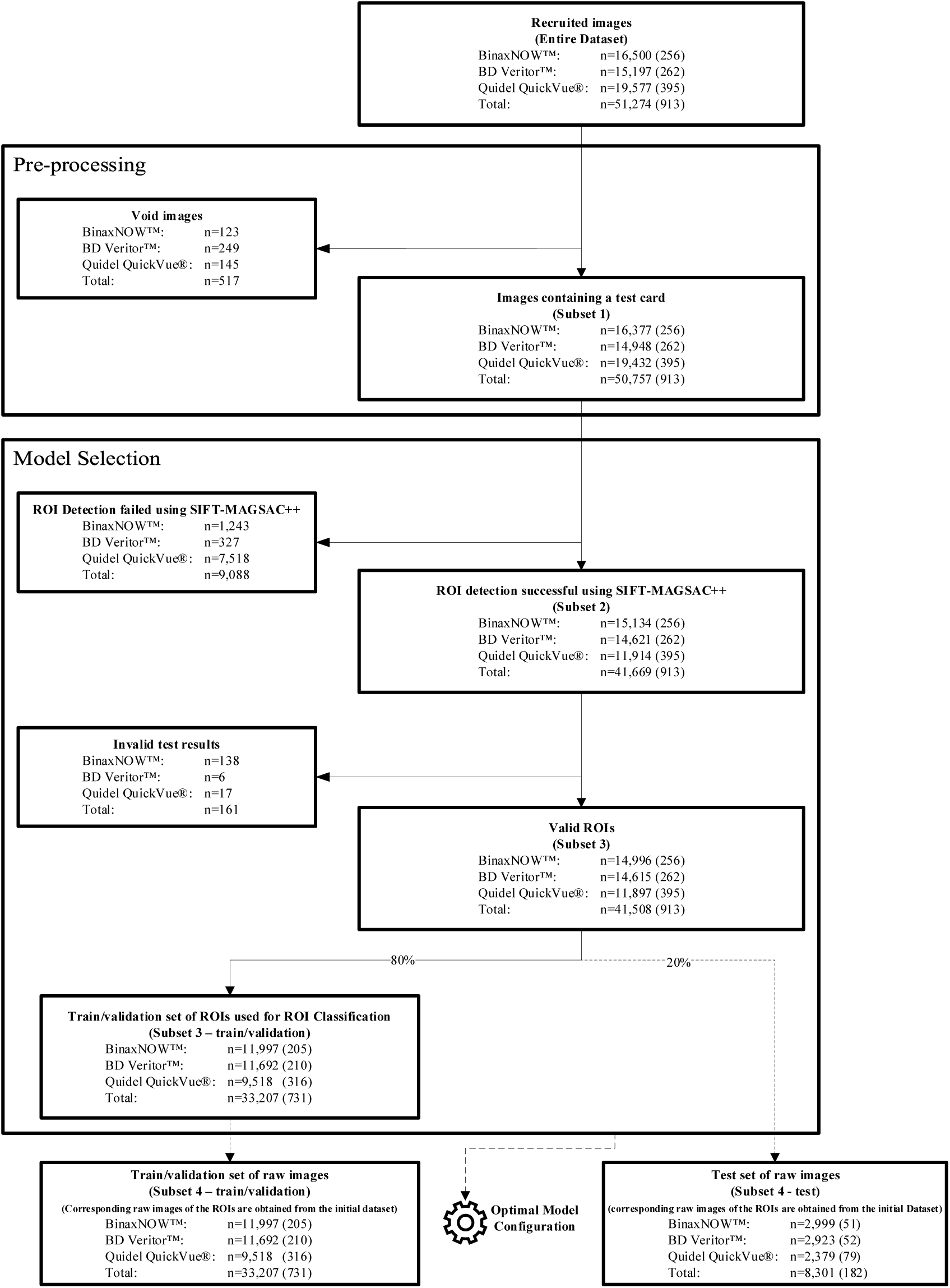
Flowchart of dataset distribution through study. The number in parentheses indicates the total number of positive samples.

Subset 2 consisted of images with successfully cropped Regions of Interest (ROIs) and was utilized for invalid result detection. To ensure the inclusion of all positive samples, some ROIs from positive samples that were not initially detected by the algorithm were manually added to Subset 2. Invalid test results were then removed, yielding Subset 3. Subset 3 was subsequently divided into train/validation and test sets randomely.

Subset 3-train/validation was employed for training purposes and for comparing different configurations of the ROI classification module. Additionally, raw images corresponding to Subset 3-train/validation was utilized to create Subset 4-train/validation. The purpose of this division was to exclude void or invalid samples from the train/validation and test sets. For more detailed information regarding the dataset distribution and study workflow, Table S2 and Fig. S6 are provided.

### Test Card Classification and ROI Detection

We evaluated the three image-matching algorithms on 1,000 inquiry images randomly sampled from subset 1 for each test card. The average number of inliers and the detection rate are shown in Table S3. The quantity of inliers serves as an indicator of similarity between the query image and the corresponding test card template (Fig. S1). We assessed the performance of three feature detection methods (SIFT, SuperPoint, ORB-BEBLID) followed by three feature matchers (FLANN, SuperGlue, and Brute Force) respectively, and three outlier removal techniques (GCRANSAC, MAGSAC++, DEGENSAC) to identify the most effective combination for ROI detection [12] [19]. The SuperPoint + SuperGlue (SP+SG) method achieved the highest performance in ROI extraction, especially with the help of DEGENSAC [20] outlier removal, and was able to detect the region of interest in 88.63% of images in total (Table S3), with Quidel QuickVue® having a lower detection rate in comparison to 93.7% and 94.7% for BinaxNOW™ and BD Veritor™, respectively. Even though the number of inliers for the BinaxNOW™ test card was greater than that of the other two, it could not be detected as effectively as the BD Veritor™ test cards. Among those images where ROI was detected, we did not observe any misclassification. The decision to use ORB+BEBLID+DEGENSAC was based on speed and computational cost. The selected method does not require GPU and can be deployed on limited-power devices such as smartphones to be run in real-time.

### Invalid Result Detection

A 5-fold cross-validation approach was employed to evaluate the performance of two models, namely the Siamese network and BiT network, in detecting invalid results within the dataset. Subset 2 was specifically used for this task, as it involved a binary classification of valid versus invalid results. The findings revealed that the BinaxNOW™ test exhibited the highest prevalence of invalid results, with a rate of 0.91%. In contrast, the Quidel QuickVue® and BD Veritor™ tests demonstrated lower rates of invalid results, at 0.14% and 0.04%, respectively. The performance of developed Siamese network was slightly better than the BiT network, and the invalid classification performance was the highest among the BD Veritor™ test cards compared to other tests (Table S4). According to our evaluation, the Siamese network was able to detect 96.8% of invalid samples in BinaxNOW™, whereas BD Veritor™ and Quidel QuickVue®, achieved detection rates of 100% and 95.7%, respectively. Examples of the detected invalid test results are provided in Fig. S7. The majority of strips classified as invalid were unused rather than truly faulty, as indicated in Table S5. Although our study did not have a sufficient number of actual invalid samples to test the module after model selection, the presence of this module in the deployment is crucial because void test results may be interpreted as negative if not eliminated.

### ROI Classification and Test Result Determination

Two networks, ResNet50 V2 and Big Transfer (BiT), were subjected to benchmark for model selection, utilizing 5-fold cross-validation on subset 3-train/validation. ResNet50 V2 serves as the backbone architecture for the BiT model. Both networks possess similar complexities and an almost equal number of parameters. However, BiT employs a specific fine-tuning technique that is well-suited for few-shot learning and unbalanced datasets, as described in reference [17].

The Big Transfer (BiT) network demonstrated higher performance in classifying the results of BD Veritor™ and Quidel QuikVue tests, achieving sensitivities of 97.3% and 93.2%, respectively (refer to Table 1). Notably, BD Veritor™ test cards exhibited higher accuracy in classification, while the accuracy for Abbott BinaxNOW™ results was comparatively lower at a sensitivity of 82.3%. This disparity can be attributed to incorrect labeling in the reference database. Fig. S8 visually presents all instances of false-negative samples observed in the Abbott BinaxNOW™ test cards. The majority of these false negatives involve faint or absent test lines, indicating the possibility of either misclassification by the study coordinators or the presence of a faint positive line that is difficult to detect through images. All misclassified samples are included in the Figs S8-13 for all test cards.

**Table 1.** ROI Classification Module Performance using 5-fold cross-validation. All metrics represent the average of the results for the five folds. AUC indicates the area under the ROC curve and was used as a measure of model evaluation.

| # | Base Model | Test Card | No. of Samples | TP | TN | FP | FN | AUC (std. dev.) | Sensitivity (% [95% CI]) | Specificity (% [95% CI]) |
| --- | --- | --- | --- | --- | --- | --- | --- | --- | --- | --- |
| 1 | Big Transfer (m-r50x1) | BinaxNOW™ | 2399 | 34 | 2351 | 7 | 7 | 0.9619 ± 0.0188 | 82.3 (71.8-92.8) | 99.7(99.5-99.9) |
|  |  | BD Veritor™ | 2338 | 41 | 2291 | 5 | 1 | 0.9874 ± 0.0059 | 97.3 (94.3-100.0) | 99.8 (99.6-99.9) |
|  |  | Quidel QuickVue® | 1904 | 59 | 1834 | 7 | 4 | 0.9852 ± 0.0146 | 93.2 (80.6-100.0) | 99.6 (99.4-99.9) |
| 2 | ResNet 50v2 | BinaxNOW™ | 2399 | 29 | 2356 | 2 | 12 | 0.9105 ± 0.0274 | 70.0 (52.7-87.2) | 99.9 (99.8-100.0) |
|  |  | BD Veritor™ | 2338 | 39 | 2289 | 7 | 3 | 0.9731 ± 0.0145 | 92.3 (87.0-97.6) | 99.7 (99.5-99.9) |
|  |  | Quidel QuickVue® | 1904 | 55 | 1835 | 6 | 8 | 0.9681 ± 0.0265 | 87.6 (73.3-100.0) | 99.7 (99.5-99.9) |

### Model Evaluation

For model evaluation, the system was trained with optimal configuration, found in previous steps, using Subset 4-train/validation. This was found to be ORB-BEBLID with DEGENSAC outlier removal, and BiT for ROI Classification. The final classification system categorized readable inputs into positive, negative, and uncertain, utilizing a threshold moving with F-score and G-mean to improve precision. The reader is encouraged to refer to the Supplementary Methods section for additional details. The F-score and G-mean were calculated based on the Subset 4-train/validation data and applied to the Subset 4-test set. The uncertain category was determined based on the area between the calculated thresholds. It is noteworthy to mention that the following results were obtained excluding the unreadable samples.

There was a substantial improvement in precision from 0.841 to 0.946 for the Abbott BinaxNOW™ test cards, with a minor decline in sensitivity and specificity when compared to a hard single threshold which is 0.5 in binary classification (Fig. 3a, b). It is noteworthy that the proportion of uncertain data classified by the models is quite low, with values of 0.012, 0.013, and 0.005 for the Abbott BinaxNOW™, Quidel QuickVue®, and BD Veritor™ test cards, respectively. Moreover, the precision of the participants was 0.850 for the same set. Additionally, the sensitivity and specificity of our method were 0.761 and 0.989, respectively, compared to participants who achieved 0.739 and 0.993, respectively, on the Abbott BinaxNOW™ test cards (Fig. S14a, b).

**Fig. 3.**
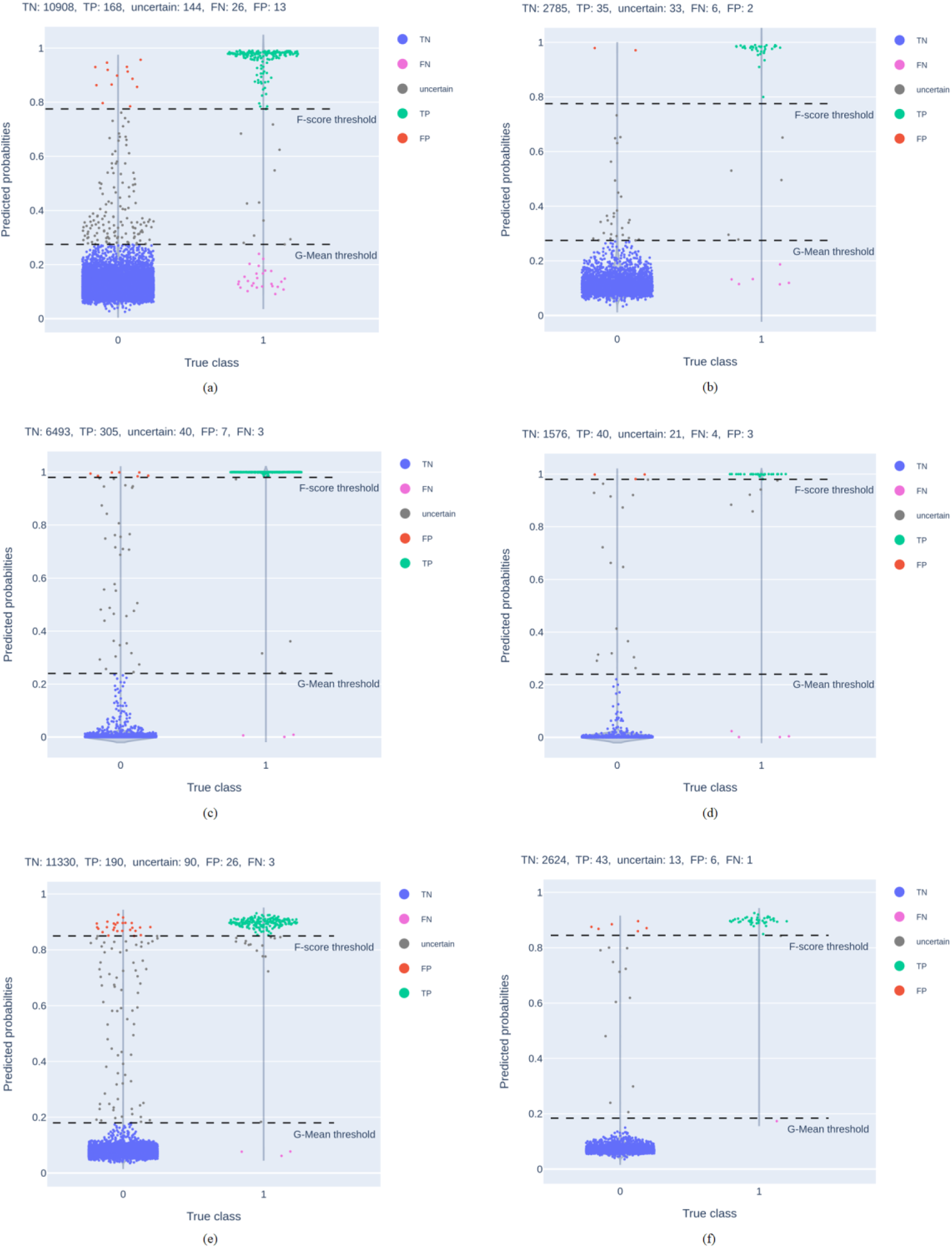
Probability plots of All test cards. **a, b** Probability plots of training/validation data and test data of the Abbott BinaxNOW™ test cards, respectively. **c, d** Probability plots of training/validation data and test data of Quidel QuickVue® test cards, respectively. **e, f** Probability plots of training/validation data and test data of BD Veritor™ test cards, respectively. For all test cards, F-score and G-mean are calculated using the training/validation data and applied to the test set. A true class of 1 indicates positive, and 0 indicates negative. Abbreviations include TN (True Negative), TP (True Positive), FP (False Positive), FN (False Negative).

For Quidel QuikVue test cards, the proposed method achieved a precision of 0.930, compared to a precision of 0.808 by participants’ interpretation (Fig. 3c, d; Fig. S14c, d). Furthermore, the proposed system demonstrated a sensitivity of 0.816 and a specificity of 0.988, whereas the participants achieved 0.775 and 0.993. These results highlight that the proposed method not only increased sensitivity but also significantly improved precision.

The BD Veritor™ test cards had the highest success rate in the classification of all tested cards (Fig. 3e, f). The achieved sensitivity, specificity, and precision for this test card were 0.955, 0.993, and 0.877, respectively, and no false negatives were observed (Fig. S14e).

## DISCUSSION

In this study, we sought to develop an automated image processing pipeline to accurately and precisely interpret the real-world images of three different COVID-19 rapid antigen tests taken by participants in their home environment. The model described in this manuscript demonstrated high precision in classifying different types of test cards, with machine interpretation surpassing human participants in terms of precision and sensitivity. However, we found heterogeneity in the performance of the model across different tests. Below, we describe the implications of our findings and important considerations regulatory agencies should pay attention to regarding the design of the test card, which could facilitate or impede the development of a universal, test-agnostic machine reader. We organize our discussion in four domains: a) the importance of real-world data, b) challenges associated with defining ground truth, c) technological constraints, and d) a path for universal machine reader.

Table 2 presents a comparative analysis of the findings from various studies on the interpretation of COVID-19 rapid antigen tests. As we observed, prior results indicate that employing multiple networks is a preferred strategy to achieve superior performance in automated COVID-19 rapid antigen test interpretation. Further, most previous studies utilized specific protocols to acquire images under ideal conditions. For instance, Peláez et al. [21] achieved excellent accuracy (100%) by utilizing an AI-based model on Panbio™ test cards. However, experienced healthcare professionals were responsible for image acquisition in hospitals using an App that ensured proper and standardized positioning of the device. Another study [9] also reported high sensitivity (0.989) and specificity (0.997) by utilizing fixed illumination. In contrast, our image database was collected from testers in their homes, with little guidance or instruction, which more closely represents the real-world use case. Additionally, most previous test readers analyzed just one type of rapid antigen test, as opposed to three analyzed by our study. We showed the variability and different challenges presented by various test types, including challenges in ROI detection for Quidel QuickVue® and low specificity for Abbott BinaxNOW™. As there are many rapid antigen tests currently on the market, it is important for any automatic reader to be versatile in its ability to detect results from multiple test types.

**Table 2.** Comparative Analysis of Similar Studies on COVID-19 Rapid Antigen Test Interpretation. Abbreviations include RT-PCR (Reverse transcriptase polymerase chain reaction), LFA (Lateral flow assay), LFIA (Lateral flow immunoassay), CNN (Convolutional Neural Network), and AI (Artificial Intelligence). Results from this study were gathered from the Model selection part for a fair comparison with other studies which considered the problem as a simple binary classification with emphasis on sensitivity and specificity.

| Study | Test type | Test card's brand | Method | with a certain protocol to acquire the image (orientation and/or pre-quality check) | ROI detection rate (%) | Invalid result detection sensitivity (%) | Ground truth | Sensitivity (% [95% CI]) | Specificity (% [95% CI]) | AUC | Size of the test set |
| --- | --- | --- | --- | --- | --- | --- | --- | --- | --- | --- | --- |
| Banathy et al. [22] Sub-study 1 | LFA | Innova LFD | Series of networks including GAN, CNN, etc. | Yes | 95.9 | 83.7 | Expert read | 97.6 (93.2-99.5) | 99.95 (99.9-99.9) | ~1 | 46,541-315+ |
| Banathy et al. [22] Sub-study 2 | LFA | Innova LFD | Series of networks including GAN, CNN, etc. | No | 92.6 | 68.4 | Expert read | 100 (90.2-100) | 99.4 (99.3-99.4) | ~1 | 56,491-177+ |
| Richardson et al. [23] | LFA | Not specified | AI-based algorithm by Exa Health, Inc. | Yes | Not studied | Not studied | RT-PCR | 86.2 (73.6-98.8) | 94.3 (88.8-99.7) | Not specified | 70-29+ |
| Wang et al. [8] | LFA | An LFA developed by GH Labs | Peak detection using image processing | Yes | Not studied | Not studied | Expert read | Not specified | 100.0 | 0.99 | 155 total |
| Peláez et al. [21] | LFA | Panbio™ by Abbott | AI-based algorithm by SpotLab | Yes | Not studied | Not studied | Expert read | 100.0 | 100.0 | 1.000 | 58-30+ |
| Wong et al. [7] | LFIA | Fortress Diagnostics | Series of networks including CNN | No | 81.3 | 91.7 (76.0-99.0) | Expert read | 90.1 (83.7-95.0) | 98.7 (96.8-99.9) | Not specified | 158-79+ |
| Mendels et al. [9] | LFIA | Trained on 10 brands | Image matching, CNN | Yes | Not studied | Not studied | Expert read | 98.9 (98.4-98.9) | 99.7 (99.4-99.9) | Not specified | 1,703-1,641+ |
| RADx-Tech CSC (BiT) | LFA | BinaxNOW™ by Abbott | Image matching, Siamese, CNN | No | 93.3 | 96.8 (91.0-100.0) | Expert read | 82.3 (72.7-91.8) | 99.7 (99.5-99.9) | 0.9619 | 2948-51+ |
| RADx-Tech CSC (BiT) | LFA | BD Veritor™ | Image matching, Siamese, CNN | Yes | 98.6 | 100.0 (100.0) | Application read | 97.3 (94.3-100.0) | 99.8 (99.6-99.9) | 0.9874 | 2858-52+ |
| RADx-Tech CSC (BiT) | LFA | Quidel QuickVue® | Image matching, Siamese, CNN | No | 74.0 | 95.7 (88.3-100.0) | Expert read | 93.2 (80.6-100.0) | 99.6 (99.4-99.9) | 0.9852 | 2287-79+ |

It is worth noting that previous studies in this field have not extensively addressed the issue of invalid test results. However, our proposed method demonstrates that training the model with a limited number of invalid test samples can effectively detect such invalid results with high accuracy. In the case of BinaxNOW™ devices, we identified four distinct categories of invalid test results, and our proposed method achieved a sensitivity of 96.8% in detecting these invalid cases. Additionally, we calculated the prevalence rates for each type of invalid test, which can aid researchers and manufacturers in understanding the underlying factors contributing to invalid results on a broader scale of usage (refer to Table S5). Our findings underscore the importance of incorporating an invalid result detection mechanism within a universal machine reader. Without such detection, the majority of invalid samples are likely to be misclassified as negative results. The comprehensive approach taken in our study to address the issue of invalid test results constitutes a unique contribution to the existing literature. However, further research is required to evaluate the performance of the invalid result detection module using a larger sample size in order to validate its efficacy.

In the studies related to ROI detection, Banathy et al. [22] achieved an outstanding detection rate of 92.63% when users were asked to submit photos of test cards. The high detection rate could be attributed to various factors such as the demographic characteristics and education of participants, instructions provided, device properties, and the performance of the detection algorithm. However, their algorithm was not able to handle rotated pictures and rejected them as unreadable, indicating the importance of the rotation-invariant property of the ROI detection algorithm. Wong et al. [7] used a 2D CNN based on the U-Net architecture and achieved a detection rate of 81.3% among over 728,000 submitted images. In our study, we investigated different methods for ROI detection and evaluated the impact of device design on the performance of the automatic interpretation system. Using SuperPoint+SuperGlue, we achieved detection rates of 93.3% and 74% for BinaxNOW™ and QuickVue® devices, respectively. However, a comparison of these results with the BD Veritor™ detection rate of 98.6% highlights the impact of test card properties and pre-quality checks that have been done on BD Veritor™ images. The diminished detection efficacy observed in QuickVue® test cards can be attributed to the variability in the visual features of the test strip, leading to a degradation in image-matching precision. Furthermore, it is advisable for manufacturers to consider incorporating visual markers, such as AprilTags, onto the test cards to enhance image-matching outcomes and reproducibility of results. Our study found that the majority of the failures in ROI detection were attributed to the device being placed too far from the camera, resulting in a decrease in the number of features to match. To mitigate this issue, visual guidance during image capture can be useful. In real-life automated systems, ROI detection is critical, and investigating different non-image matching algorithms may help to improve their reliability and accuracy. Finally, it should be noted that the performance of the system described in this study surpasses that of human interpretations, but this superiority is contingent upon the image being captured under favorable conditions. Thus, it is recommended that prospective companies offering similar reader applications incorporate additional guidance for image capture to ensure a robust and universally applicable system.

Our study is not exempt from limitations. While image matching proves effective in handling scale and rotation variations, it presents significant computational expenses and time constraints. Additionally, our system requires a minimum number of inliers to classify the submitted image as readable, leading to potential rejections despite accurate detection of the ROI due to a low number of inliers. Lastly, the ground truth for the model was the “expert read” of the rapid antigen test by study coordinators. While this should be less prone to user bias than reading one’s test, this is still subject to user errors and limitations inherent to human visual perception. To address these concerns, we aim to evaluate our model against paired RT-PCR results and results from other COVID-19 diagnostics, to further enhance and train the model.

## CONCLUSION

We proposed an automatic decision system aimed at interpreting Ag-RDTs using image matching and deep learning techniques. We employed multiple image-matching algorithms and carried out an ablation study using real participant-submitted images. Our findings indicate that all three image-matching algorithms performed comparably. However, we emphasized on exploration of invalid result detection in the current approach, in contrast to previous methods in this field. The current study sheds light on the challenges, limitations, and corresponding possible solutions on the path to deploying a fully automatic, robust, and precise diagnostic system. Overall, our results suggest that this pipeline could serve as a valuable tool in at-home settings for the interpretation of lateral flow assays, facilitating the precise and reliable detection of COVID-positive individuals. This pipeline could not only be used to reduce bias in result interpretation, but it could also address accessibility issues for many people who suffer from visual or cognitive impairments that may interfere with accurate self-interpretation of over-the-counter tests.

## Data Availability

All data produced in the present study are available upon reasonable request to the authors

## CODE AVAILABILITY

