## Supplementary material for "Automated Classification of At-home SARS-CoV-2 Lateral Flow Assay Test Results using Image Matching and Transfer Learning: multiple-pipeline study": Supplemetal Materials

Supplementary Figures:

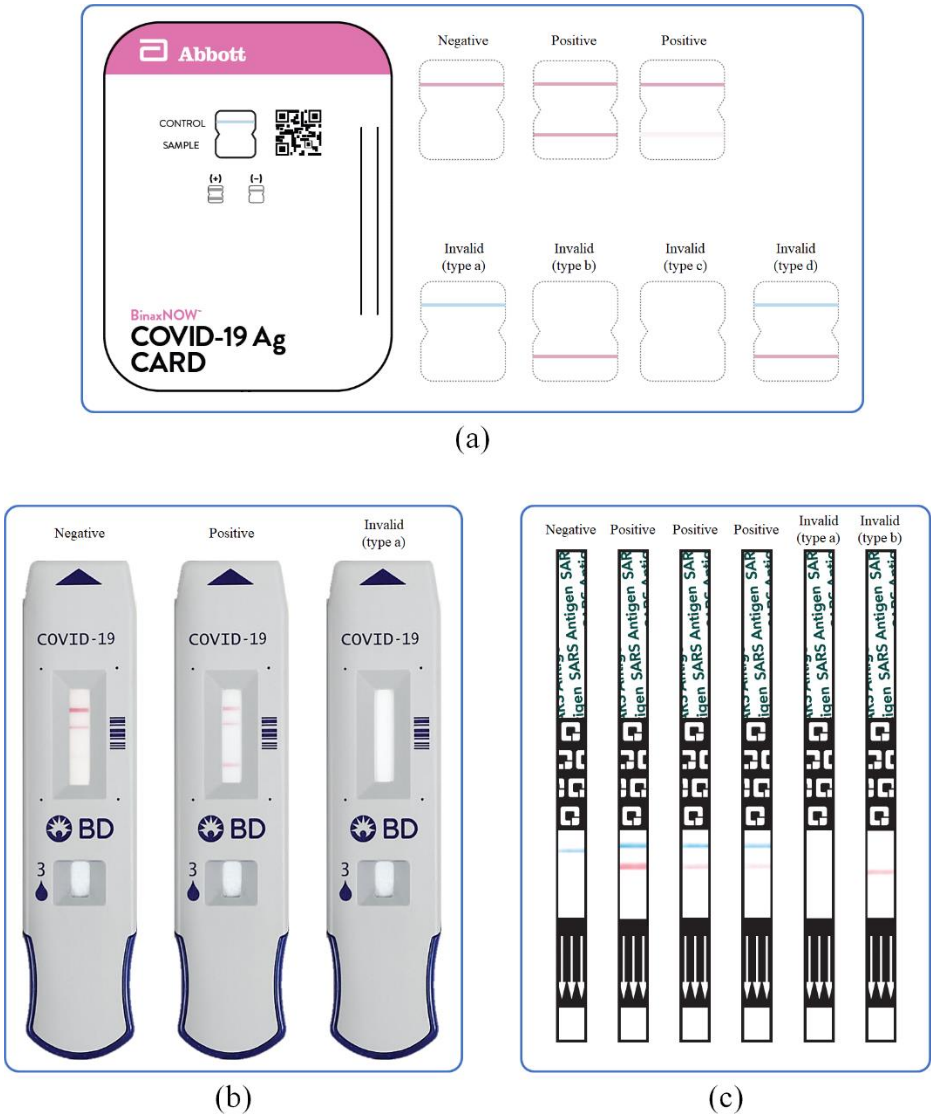

**Fig. S1 Test cards used in the study. a** Abbott BinaxNOW™ test card and its test results examples. If there is a red control line in the result window, the test is done correctly, and the result can be either negative or positive. If the red control line is absent, the result is interpreted as invalid. The test card on the left is unused. **b** Test result examples of BD Veritor™ assay. Since the interpretation criteria are not published publicly, we consider only unused cards as invalid test results, and we only have the test result samples and their corresponding labels provided by the "Scanwell Health" app. **c** Test result examples of Quidel QuickVue® test strip. The unused test card is as same as the invalid type a.

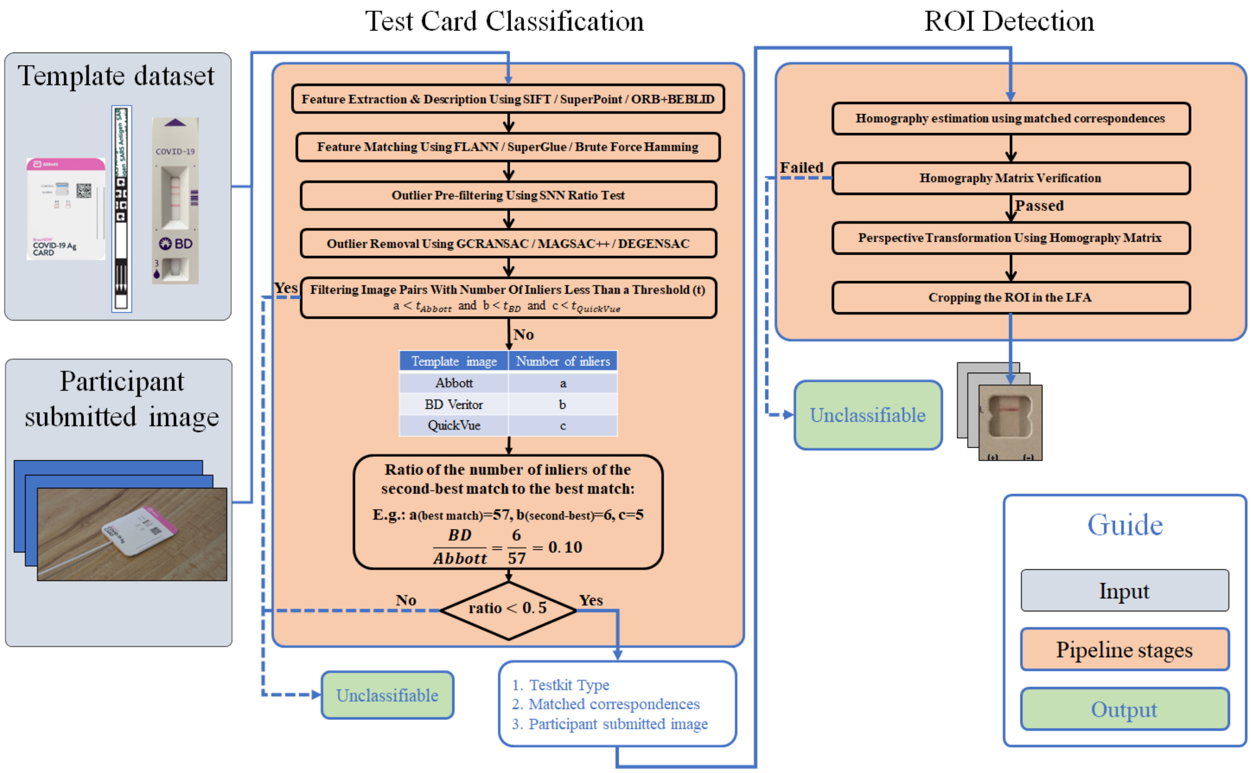

**Fig. S2 The detailed workflow of the Test Card Classification and ROI Detection modules.**

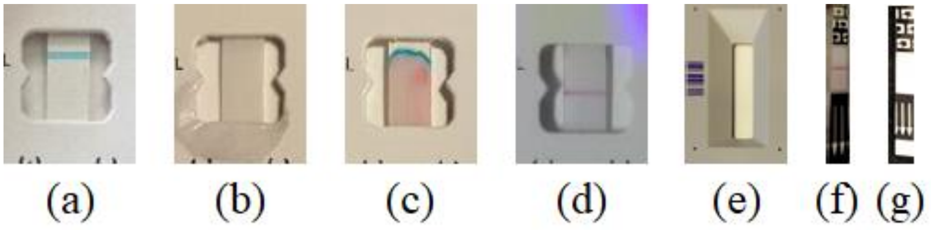

**Fig. S3 All types of actual invalid test results. a, b, c, d** Four types of invalid results of Abbott BinaxNOW test card. **e** Invalid test result of BD Veritor test card. **f, g** Two types of invalid results of Quidel QuickVue test card.

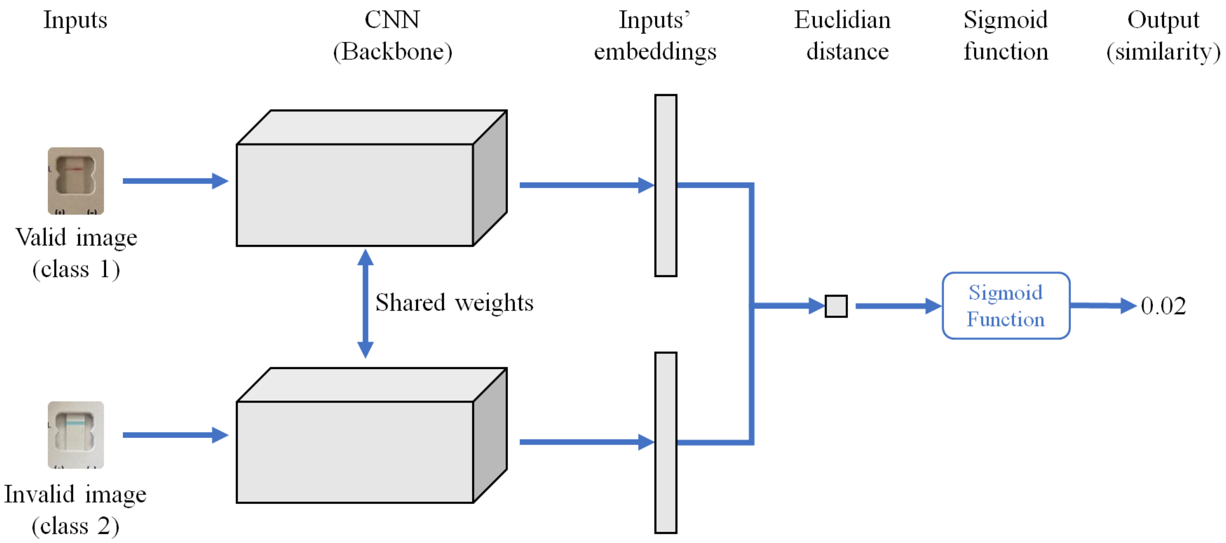

**Fig. S4 Proposed Siamese neural network for Invalid Detection module.** It comprises a twin CNN with shared weights. It was developed such that in the training phase, each image was paired once with the same class image and once with a different class image. This helps the network to learn faster with fewer samples. For instance, in this figure, first input is belonging to class 1 (void images) and second input is belonging to class 2 (invalid images). Therefore, the similarity is very close to zero.

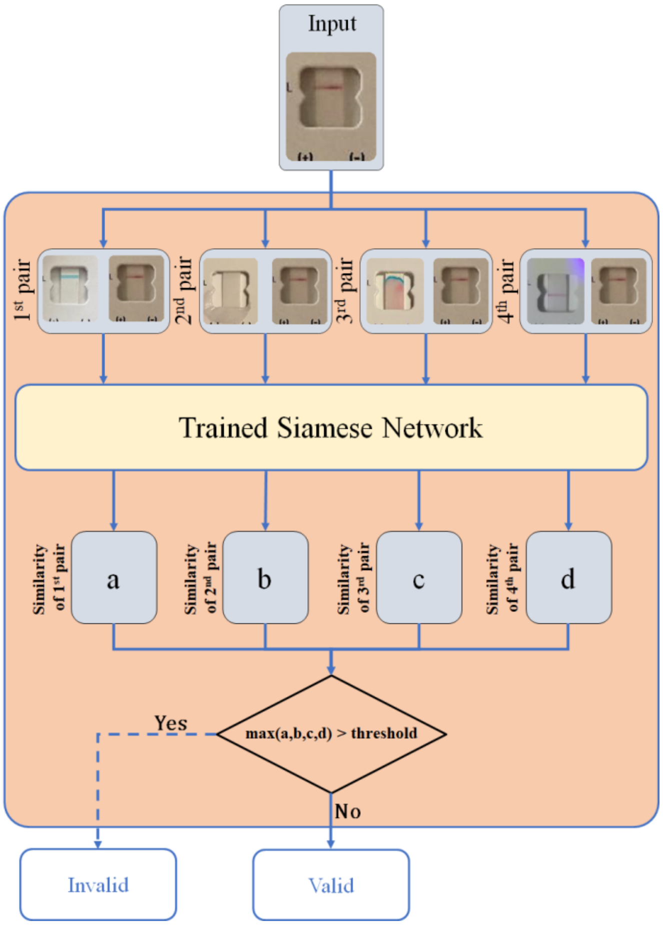

**Fig. S5 Flowchart of the Invalid Detection module.** Each input forms four pairs with different invalid templates. The invalid templates are gathered from the user instruction of each test card for the sake of representation.

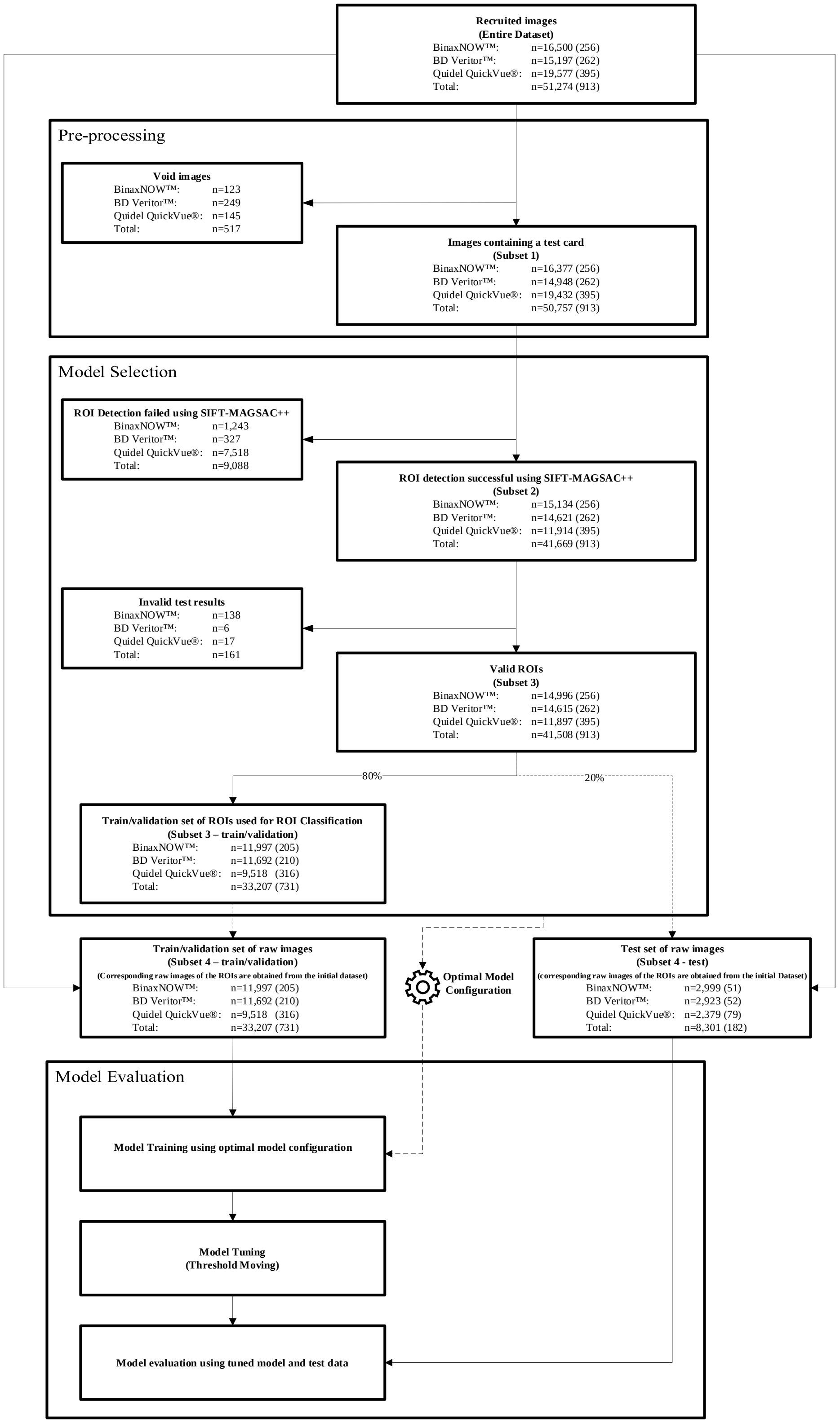

**Fig. S6 Study workflow.** The study workflow consisted of a thorough investigation of various model configurations using subset 3. After determining the optimal configuration, the model was trained using training/validation data and fine-tuned using threshold moving. Finally, the selected model was evaluated using a test set.

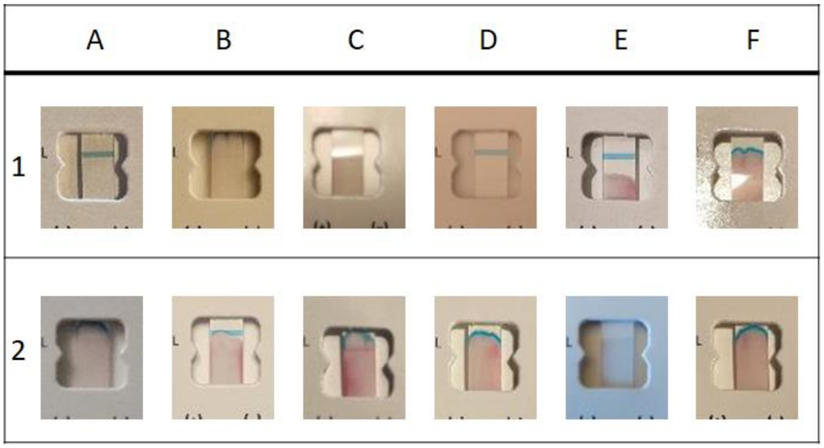

**Fig. S7 Some examples of Abbott BinaxNOW invalid test results detected by the Invalid detection running on the whole dataset.**

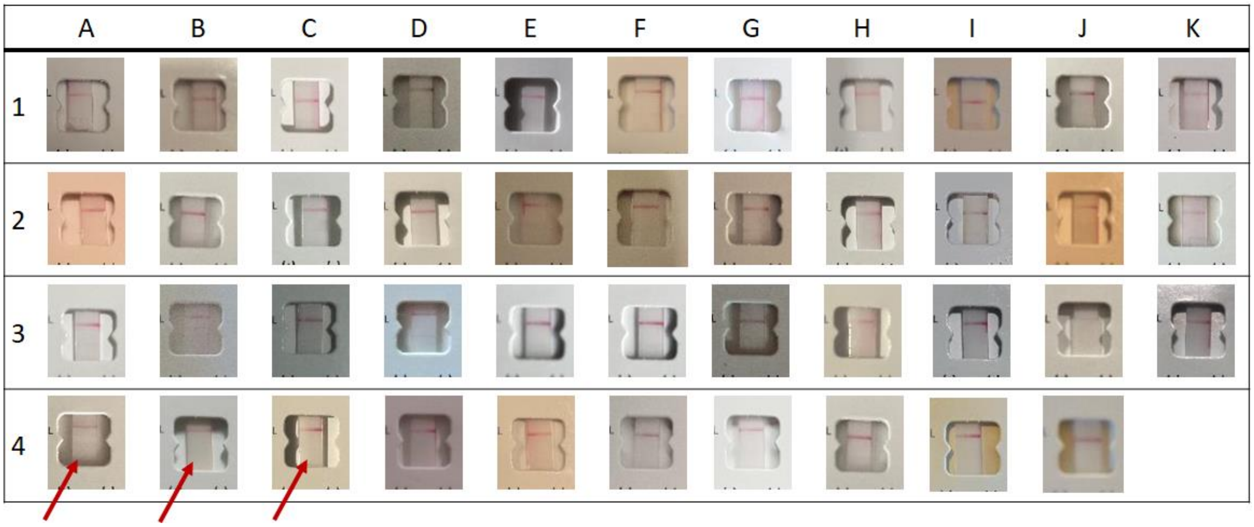

**Fig. S8 All false negative samples for Abbott BinaxNOW™ test cards.** They are gathered using the Big Transfer model. As can be seen, most of the tests labeled as “positive” by participants either lack a test line or the test line is barely visible. For instance, in row 4, columns A, B, and C depict samples lacking a test line, as indicated by arrows.

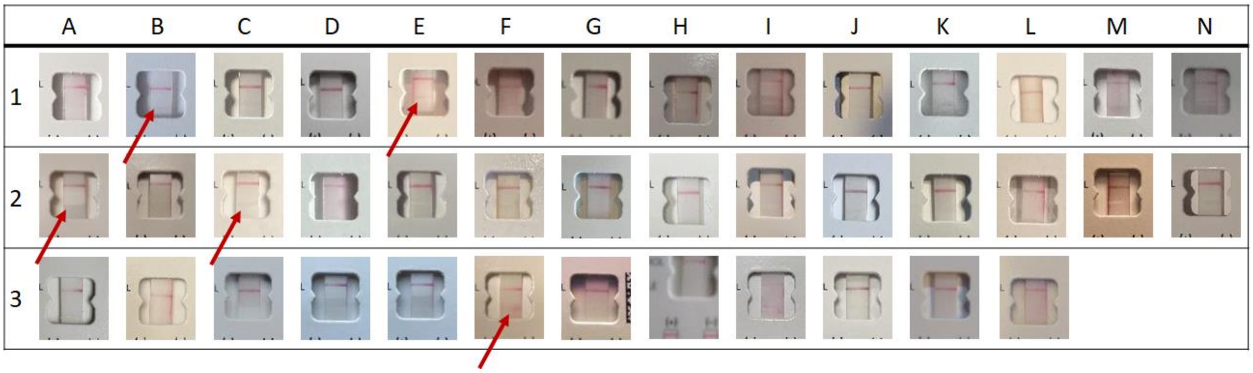

**Fig. S9 All false positive samples for Abbott BinaxNOW™ test cards.** They were collected from all five test folds using the Big Transfer model. As demenstrated in some cases a faint test line is visible that are shown with a red arrow.

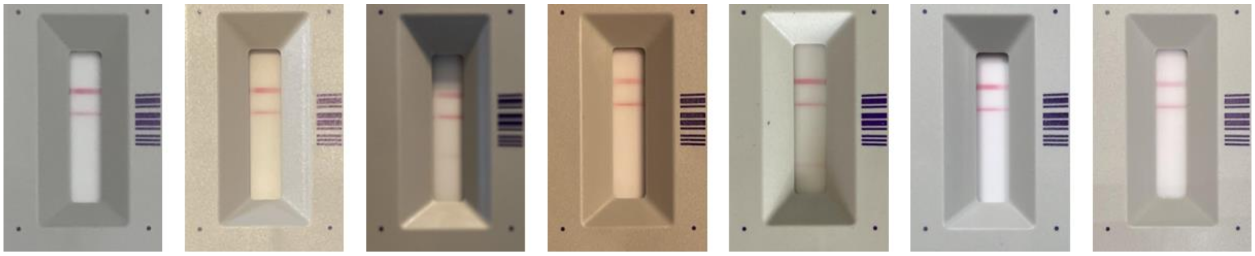

(a) (b) (c) (d) (e) (f) (e)

**Fig. S10 All false negative samples of BD Veritor test cards.** They were gathered from all five test folds using the Big Transfer model. As can be seen most of them either lack the third line although they are labeled as positive, or the test line is extremely weak. It is worth mentioning that interpretation instruction is not provided publicly by the manufacturer.

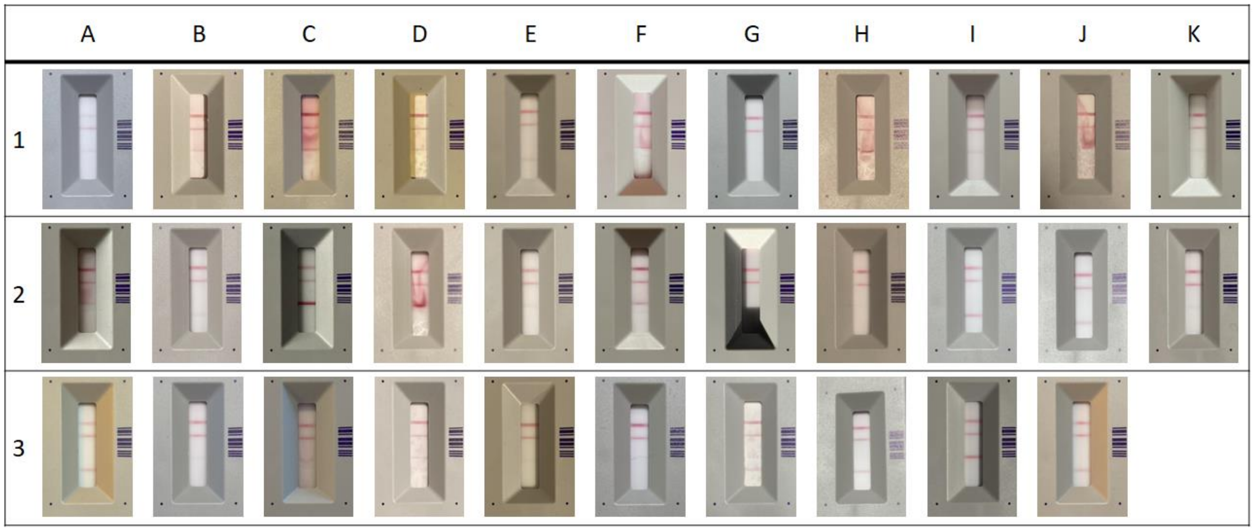

**Fig. S11 All false positive samples for BD Veritor test cards.** They were collected from all five test folds using the Big Transfer model. The results indicate that the majority of these samples have a third line, which based on our observations in the dataset is an indicator of a positive test result. Additionally, some samples have a third line diminished in their area. Notably, the interpretation instructions for BD test cards are not publicly available from the manufacturer.

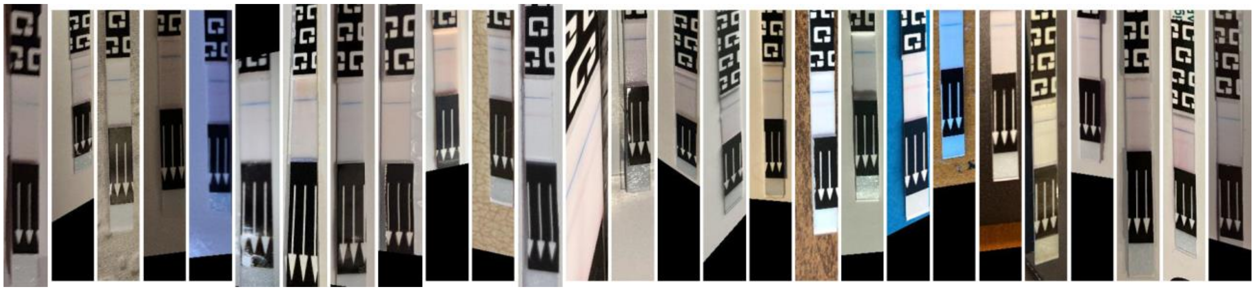

**Fig. S12 All false negative samples of Quidel QuickVue test cards.** They were collected from all five test folds were processed using the Big Transfer model. The results indicate that in most cases, the test line was either barely detectable or not visible at all.

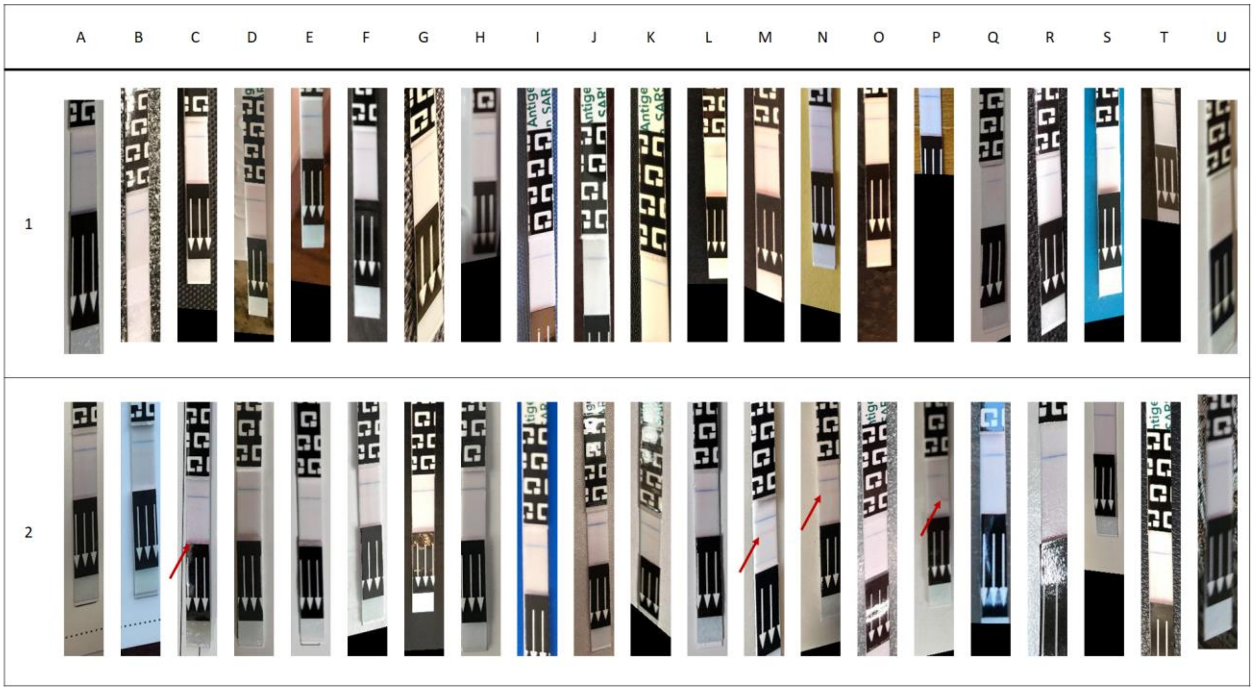

**Fig. S13 All false positive samples of Quidel QuickVue test cards.** They were collected from all five test folds were processed using the Big Transfer model. The results reveal that in certain cases such as M2, N2, a very faint test line is visible. Additionally, in some cases like C2, there is a red line that appears at the bottom of the strip, which may have misled the classifier. Another infrequent case seen in P2 involves a diminished test line that still exists and could potentially mislead the classifier.

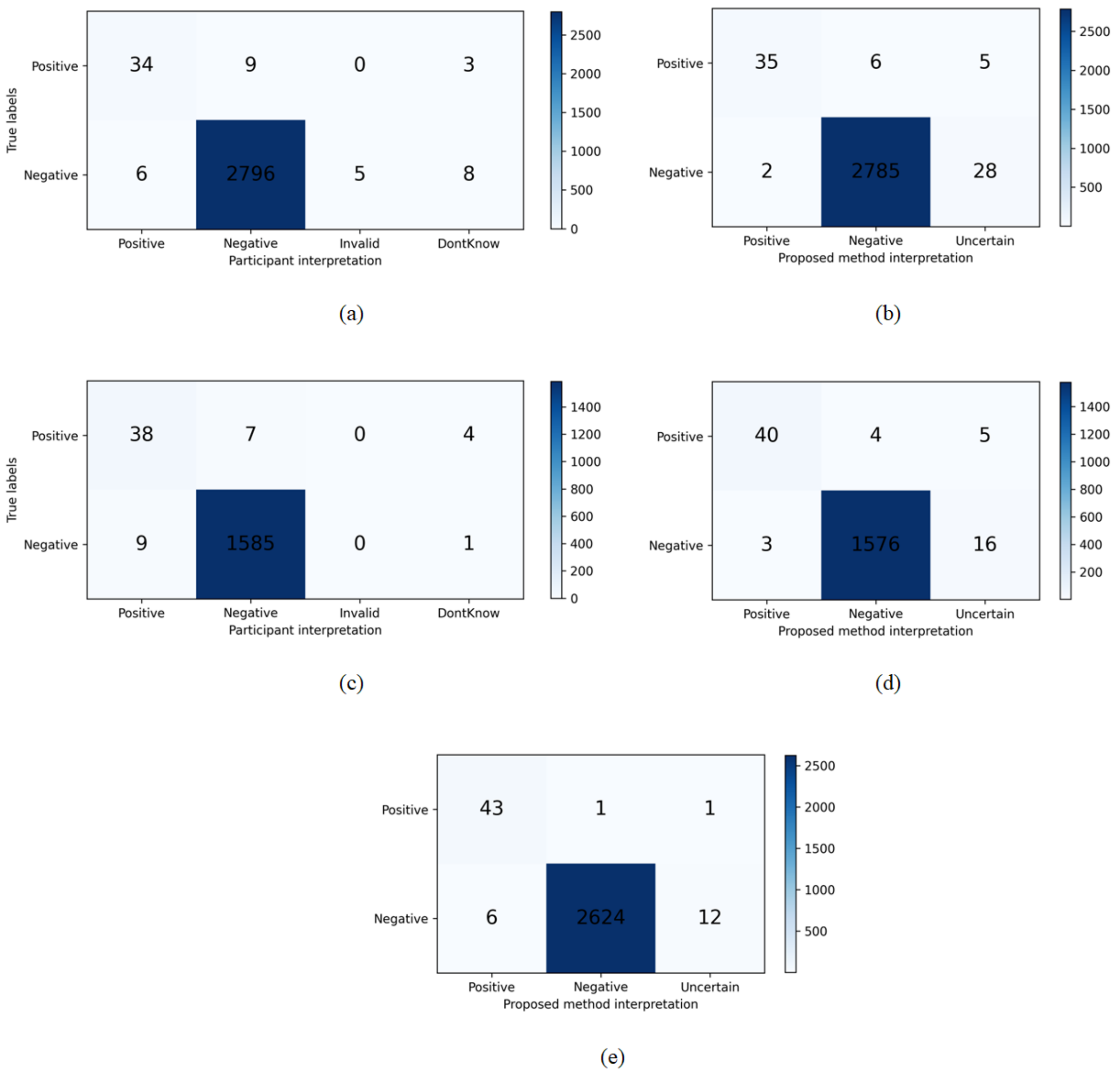

**Fig. S14 Confusion matrices of participants vs proposed method. a, b** The interpretation of the participants on the test set and the proposed method on the same set for Abbott BinaxNOW™ test cards. **c, d** The interpretation of the participants and the proposed method on the test set for Quidel QuickVue® test cards. **e** The interpretation of the proposed method on the test set for BD Veritor™ test cards. BD Veritor™ test card is not designed to be interpreted by the participants, instead an associated application is designed to do so.

### Supplementary Tables:

**Table S1 Invalid Detection Module Configuration.** Note that optimal value is obtained using GridSearch with the specified parameters.

| Method | Parameter | Optimal value (in most cases) | Values used in hyperparameter tuning using GridSearch |
|---|---|---|---|
| Proposed Siamese Network | Batch size | Abbott:32 , BD:8, QuickVue:8 | 8, 16, 32 |
| | Optimizer | SGD | - |
| | Learning rate | ExponentialDecay<br>decay_steps=3000<br>decay_rate=0.96 | -<br>- |
| | Backbone weights | ImageNet-21k | - |
| | Backbone name | m-r50x1 | - |
| | Loss | ContrastiveLoss | - |
| | SGD momentum | 0.9 | 0.9 |
| | Epochs | 5 | Early stopping is used to find the best epoch |
| Big Transfer | Batch size | Abbott:64, BD:32, QuickVue:8 | 8, 16, 32, 64, 128 |
| | Optimizer | SGD | - |
| | Learning rate | piecewise constant decay schedule as mentioned in [6] | - |
| | Backbone weights | ImageNet-21k | - |
| | Backbone name | m-r50x1 | - |
| | SGD momentum | 0.9 | 0.9 |
| | Schedule length | 500 | - |
| | Class weight | 1.0:10.0 | 1:1, 1:2, 1:4, 1:6, 1:8, 1:10 |

**Table S2 Dataset properties.** Positive tests indicate the number of positive samples among all.

| Test card | Category | Submitted photo (Entire dataset) | The photo contains a test card (Subset 1) | ROI Detection successful using SIFT-MAGSAC++ (Subset 2) | Valid Result used for ROI classification (Subset 3) |
|---|---|---|---|---|---|
| BinaxNOW™ | All | 16,500 | 16,377 | 15,134 | 14,996 |
| | Participants | 2,238 | 2,232 | 2,221 | 2,218 |
| | Positive tests | 256 | 256 | 256 | 256 |
| BD Veritor | All | 15,197 | 14,948 | 14,621 | 14,615 |
| | Participants | 2,131 | 2,113 | 2,108 | 2,108 |
| | Positive tests | 262 | 262 | 262 | 258 |
| Quidel QuikVue | All | 19,577 | 19,432 | 11,914 | 11,897 |
| | Participants | 2,583 | 2,569 | 2,392 | 2,392 |
| | Positive tests | 395 | 395 | 395 | 395 |
| Total | All | 51,274 | 50,757 | 41,669 | 41,508 |
| | Participants | 6,952 | 6,914 | 6,721 | 6,718 |
| | Positive tests | 877 | 877 | 877 | 877 |

**Table S3 Evaluation of Test Card Classification Module.** The detection rate demonstrates the percentage of data detectable by this module. The average number of inliers can be used as a criterion to indicate the confidence of the detected ROI.

| Method | | | Average number of inliers | | | Detection rate (%) | | | |
|---|---|---|---|---|---|---|---|---|---|
| Feature detection | Feature matcher | Outlier removal | BinaxNOW™ | BD Veritor™ | Quidel QuickVue® | BinaxNOW™ | BD Veritor™ | Quidel QuickVue® | Average |
| SIFT [1] | FLANN | GCRANSAC | 59 | 67 | 24 | 92.0 | 98.1 | 53.0 | 81.03 |
| | | MAGSAC++ | 60 | 67 | 24 | 92.2 | 98.2 | 50.7 | 80.36 |
| | | DEGENSAC | 71 | 76 | 32 | 93.8 | 99.9 | 66.8 | 86.83 |
| SuperPoint [2] | SuperGlue [3] | GCRANSAC | 99 | 77 | 28 | 92.7 | 98.4 | 67.7 | 86.26 |
| | | MAGSAC++ | 99 | 76 | 27 | 92.5 | 98.4 | 68.6 | 86.50 |
| | | DEGENSAC | 263 | 102 | 46 | 93.3 | 98.6 | 74.0 | 88.63 |
| ORB [4] & BEBLID [5] | Brute Force | GCRANSAC | 84 | 52 | 35 | 89.8 | 91.3 | 56.8 | 79.29 |
| | | MAGSAC++ | 83 | 52 | 34 | 90.8 | 91.8 | 53.7 | 78.76 |
| | | DEGENSAC | 130 | 56 | 42 | 93.7 | 94.7 | 62.9 | 83.76 |
| | | Average | 105 | 69 | 32 | 92.3 | 96.6 | 61.6 | |

**Table S4 Performance of the Invalid result detection using 5-fold cross-validation.** All metrics represent the average of the results for the five folds (test sets). In this module, true positive samples refer to invalid samples that were correctly detected as invalid, while true negative samples refer to valid samples that were correctly identified as valid. Similarly, false positive samples correspond to valid samples that were incorrectly classified as invalid, and false negative samples represent invalid samples that were misclassified as valid.

| # | Model | Test Card | No. of Samples | TP | TN | FP | FN | AUC (std. dev.) | Sensitivity (% [95% CI]) | Specificity (% [95% CI]) |
|---|---|---|---|---|---|---|---|---|---|---|
| 1 | Siamese network | BinaxNOW™ | 3036 | 35 | 2997 | 3 | 1 | 0.9880 ± 0.0142 | 96.8 (91.0-100.0) | 99.9 (99.8-100.0) |
| | | BD Veritor™ | 2933 | 10 | 2917 | 6 | 0 | 0.9949 ± 0.0000 | 100.0 (100.0) | 99.8 (99.6-99.9) |
| | | Quidel QuikVue | 2411 | 27 | 2352 | 31 | 1 | 0.9935 ± 0.0023 | 95.7 (88.3-100.0) | 98.7 (98.2-99.1) |
| 2 | BiT network | BinaxNOW™ | 3036 | 34 | 2987 | 13 | 2 | 0.9945 ± 0.0006 | 95.7 (89.0-100.0) | 99.6 (97.4-100.0) |
| | | BD Veritor™ | 2933 | 10 | 2912 | 11 | 0 | 0.9949 ± 0.0007 | 100.0 (100.0) | 99.6 (99.4-99.8) |
| | | Quidel QuikVue | 2411 | 27 | 2362 | 21 | 1 | 0.9940 ± 0.0024 | 95.8 (88.4-100.0) | 99.1 (98.8-99.5) |

**Table S5 The prevalence of each invalid type in the invalid set of data**

| | No. of cases (Prevalence rate %) | | | | |
|---|---|---|---|---|---|
| Invalid type | a | b | c | d | Total |
| BinaxNOW | 66 (47.8 %) | 18 (13.1%) | 28 (20.3%) | 26 (18.8%) | 138 |
| BD | 6 (100%) | N/A | N/A | N/A | 6 |
| QuickVue | 17 (100%) | 0 (0%) | N/A | N/A | 17 |

**Table S6 Test card Classification Module configuration**

| Method | Parameter | Value |
|---|---|---|
| SIFT | Number of features | 0 |
| | Edge threshold | 10 |
| | Number of octave layers | 3 |
| | Contrast threshold | 0.04 |
| | Sigma | 1.6 |
| | SNN threshold | 0.8 |
| | Descriptor matcher | FLANN |
| | $t_{Abbott}$, $t_{BD}$, $t_{QuickVue}$ | 10, 15, 22 |
| SuperPoint | Number of radii | 4 |
| | Keypoint threshold | 0.005 |
| | Max number of keypoints | 1024 |
| | Sinkhorn iterations | 20 |
| | Match threshold | 0.1 |
| | Descriptor matcher | SuperGlue |
| | Weights | outdoor |
| | $t_{Abbott}$, $t_{BD}$, $t_{QuickVue}$ | 10, 12, 22 |
| ORB + BEBLID | Number of features | 10000 |
| | Scale factor | 1.2 |
| | Number of levels | 8 |
| | Edge threshold | 31 |
| | First level | 0 |
| | Patch size | 31 |
| | Fast threshold | 20 |
| | Descriptor matcher | BruteForce Hamming |
| | BEBLID scale | 0.75 |
| | $t_{Abbott}$, $t_{BD}$, $t_{QuickVue}$ | 10, 9, 22 |

**Table S7 Summary of proposed Siamese network.** The input should be changed as the dimension of the ROI. Note that resnet_1 and resnet_2 have the same weights.

| | **Name** | **Type** | **Connected to** | **Output Shape** |
|---|---|---|---|---|
| **0** | input_1 | InputLayer | input_1 | [(None, 90, 75, 3)] |
| **1** | input_2 | InputLayer | input_2 | [(None, 90, 75, 3)] |
| **2** | resnet_1 | Resnet50 V2 (m-r50x1) | input_1 | (None, 2048) |
| **2** | resnet_2 | Resnet50 V2 (m-r50x1) | input_2 | (None, 2048) |
| **3** | lambda (Euclidean distance) | Euclidean distance | resnet_1 resnet_2 | (None, 1) |
| **4** | batch_normalization | BatchNormalization | lambda | (None, 1) |
| **5** | dense_4 | Dense | batch_normalization | (None, 1) |

**Table S8 ROI Classification Module configuration**

| Method | Parameter | Optimal value (in most cases) | Values used in hyperparameter tuning using GridSearch |
|---|---|---|---|
| Big Transfer | Batch size | Abbott:64, BD:32, QuickVue:8 | 8, 16, 32, 64, 128 |
| | Optimizer | SGD | - |
| | Learning rate | piecewise constant decay schedule as mentioned in [6] | - |
| | Backbone weights | ImageNet-21k | - |
| | Backbone name | m-r50x1 | - |
| | SGD momentum | 0.9 | 0.9 |
| | Schedule length | 500 | - |
| | Class weight | 1.0:10.0 | 1:1, 1:2, 1:4, 1:6, 1:8, 1:10 |
| | Epochs | 5 | Early stopping is used to find the best epoch |
| Modified ResNet 50V2 | Batch size | Abbott:64, BD:32, QuickVue:8 | 8, 16, 32, 64, 128 |
| | Optimizer | Adam | Adam, SGD |
| | Learning rate | 1e-4 | 1e-3:1e-5:5e-4 |
| | Backbone weights | ImageNet | - |
| | Backbone name | ResNet50 V2 | - |
| | Loss function | cross-entropy | - |
| | Class weight | 1.0:10.0 | 1:1, 1:2, 1:4, 1:6, 1:8, 1:10 |
| | Epochs | 5 | Early stopping is used to find the best epoch |

**Table S9 The details of modified ResNet 50V2. From row 191 to the end is added to the original architecture.**

| | Name | Type | Connected to | Output Shape |
|---|---|---|---|---|
| **0** | input_2 | InputLayer | input_2 | [(None, 224, 224, 3)] |
| **1** | conv1_pad | ZeroPadding2D | input_2 | (None, 230, 230, 3) |
| **2** | conv1_conv | Conv2D | conv1_pad | (None, 112, 112, 64) |
| **3** | pool1_pad | ZeroPadding2D | conv1_conv | (None, 114, 114, 64) |
| **4** | pool1_pool | MaxPooling2D | pool1_pad | (None, 56, 56, 64) |
| **5** | conv2_block1_preact_bn | BatchNormalization | pool1_pool | (None, 56, 56, 64) |
| **6** | conv2_block1_preact_relu | Activation | conv2_block1_preact_bn | (None, 56, 56, 64) |
| **7** | conv2_block1_1_conv | Conv2D | conv2_block1_preact_relu | (None, 56, 56, 64) |
| **8** | conv2_block1_1_bn | BatchNormalization | conv2_block1_1_conv | (None, 56, 56, 64) |
| **9** | conv2_block1_1_relu | Activation | conv2_block1_1_bn | (None, 56, 56, 64) |
| **10** | conv2_block1_2_pad | ZeroPadding2D | conv2_block1_1_relu | (None, 58, 58, 64) |
| **11** | conv2_block1_2_conv | Conv2D | conv2_block1_2_pad | (None, 56, 56, 64) |
| **12** | conv2_block1_2_bn | BatchNormalization | conv2_block1_2_conv | (None, 56, 56, 64) |
| **13** | conv2_block1_2_relu | Activation | conv2_block1_2_bn | (None, 56, 56, 64) |
| **14** | conv2_block1_0_conv | Conv2D | conv2_block1_preact_relu | (None, 56, 56, 256) |
| **15** | conv2_block1_3_conv | Conv2D | conv2_block1_2_relu | (None, 56, 56, 256) |
| **16** | conv2_block1_out | Add | conv2_block1_0_conv conv2_block1_3_conv | (None, 56, 56, 256) |
| **17** | conv2_block2_preact_bn | BatchNormalization | conv2_block1_out | (None, 56, 56, 256) |
| **18** | conv2_block2_preact_relu | Activation | conv2_block2_preact_bn | (None, 56, 56, 256) |
| **19** | conv2_block2_1_conv | Conv2D | conv2_block2_preact_relu | (None, 56, 56, 64) |
| **20** | conv2_block2_1_bn | BatchNormalization | conv2_block2_1_conv | (None, 56, 56, 64) |
| **21** | conv2_block2_1_relu | Activation | conv2_block2_1_bn | (None, 56, 56, 64) |
| **22** | conv2_block2_2_pad | ZeroPadding2D | conv2_block2_1_relu | (None, 58, 58, 64) |
| **23** | conv2_block2_2_conv | Conv2D | conv2_block2_2_pad | (None, 56, 56, 64) |
| **24** | conv2_block2_2_bn | BatchNormalization | conv2_block2_2_conv | (None, 56, 56, 64) |
| **25** | conv2_block2_2_relu | Activation | conv2_block2_2_bn | (None, 56, 56, 64) |
| **26** | conv2_block2_3_conv | Conv2D | conv2_block2_2_relu | (None, 56, 56, 256) |
| **27** | conv2_block2_out | Add | conv2_block1_out conv2_block2_3_conv | (None, 56, 56, 256) |
| **28** | conv2_block3_preact_bn | BatchNormalization | conv2_block2_out | (None, 56, 56, 256) |
| **29** | conv2_block3_preact_relu | Activation | conv2_block3_preact_bn | (None, 56, 56, 256) |
| **30** | conv2_block3_1_conv | Conv2D | conv2_block3_preact_relu | (None, 56, 56, 64) |
| **31** | conv2_block3_1_bn | BatchNormalization | conv2_block3_1_conv | (None, 56, 56, 64) |
| **32** | conv2_block3_1_relu | Activation | conv2_block3_1_bn | (None, 56, 56, 64) |
| **33** | conv2_block3_2_pad | ZeroPadding2D | conv2_block3_1_relu | (None, 58, 58, 64) |
| **34** | conv2_block3_2_conv | Conv2D | conv2_block3_2_pad | (None, 28, 28, 64) |
| **35** | conv2_block3_2_bn | BatchNormalization | conv2_block3_2_conv | (None, 28, 28, 64) |
| **36** | conv2_block3_2_relu | Activation | conv2_block3_2_bn | (None, 28, 28, 64) |
| **37** | max_pooling2d_3 | MaxPooling2D | conv2_block2_out | (None, 28, 28, 256) |
| **38** | conv2_block3_3_conv | Conv2D | conv2_block3_2_relu | (None, 28, 28, 256) |
| **39** | conv2_block3_out | Add | max_pooling2d_3 conv2_block3_3_conv | (None, 28, 28, 256) |
| **40** | conv3_block1_preact_bn | BatchNormalization | conv2_block3_out | (None, 28, 28, 256) |
| **41** | conv3_block1_preact_relu | Activation | conv3_block1_preact_bn | (None, 28, 28, 256) |
| **42** | conv3_block1_1_conv | Conv2D | conv3_block1_preact_relu | (None, 28, 28, 128) |
| **43** | conv3_block1_1_bn | BatchNormalization | conv3_block1_1_conv | (None, 28, 28, 128) |
| **44** | conv3_block1_1_relu | Activation | conv3_block1_1_bn | (None, 28, 28, 128) |
| **45** | conv3_block1_2_pad | ZeroPadding2D | conv3_block1_1_relu | (None, 30, 30, 128) |
| **46** | conv3_block1_2_conv | Conv2D | conv3_block1_2_pad | (None, 28, 28, 128) |
| **47** | conv3_block1_2_bn | BatchNormalization | conv3_block1_2_conv | (None, 28, 28, 128) |
| **48** | conv3_block1_2_relu | Activation | conv3_block1_2_bn | (None, 28, 28, 128) |
| **49** | conv3_block1_0_conv | Conv2D | conv3_block1_preact_relu | (None, 28, 28, 512) |
| **50** | conv3_block1_3_conv | Conv2D | conv3_block1_2_relu | (None, 28, 28, 512) |
| **51** | conv3_block1_out | Add | conv3_block1_0_conv conv3_block1_3_conv | (None, 28, 28, 512) |
| **52** | conv3_block2_preact_bn | BatchNormalization | conv3_block1_out | (None, 28, 28, 512) |
| **53** | conv3_block2_preact_relu | Activation | conv3_block2_preact_bn | (None, 28, 28, 512) |
| **54** | conv3_block2_1_conv | Conv2D | conv3_block2_preact_relu | (None, 28, 28, 128) |
| **55** | conv3_block2_1_bn | BatchNormalization | conv3_block2_1_conv | (None, 28, 28, 128) |
| **56** | conv3_block2_1_relu | Activation | conv3_block2_1_bn | (None, 28, 28, 128) |
| **57** | conv3_block2_2_pad | ZeroPadding2D | conv3_block2_1_relu | (None, 30, 30, 128) |
| **58** | conv3_block2_2_conv | Conv2D | conv3_block2_2_pad | (None, 28, 28, 128) |
| **59** | conv3_block2_2_bn | BatchNormalization | conv3_block2_2_conv | (None, 28, 28, 128) |
| **60** | conv3_block2_2_relu | Activation | conv3_block2_2_bn | (None, 28, 28, 128) |
| **61** | conv3_block2_3_conv | Conv2D | conv3_block2_2_relu | (None, 28, 28, 512) |
| **62** | conv3_block2_out | Add | conv3_block1_out conv3_block2_3_conv | (None, 28, 28, 512) |

| | | | | |
|---|---|---|---|---|
| **63** | conv3_block3_preact_bn | BatchNormalization | conv3_block2_out | (None, 28, 28, 512) |
| **64** | conv3_block3_preact_relu | Activation | conv3_block3_preact_bn | (None, 28, 28, 512) |
| **65** | conv3_block3_1_conv | Conv2D | conv3_block3_preact_relu | (None, 28, 28, 128) |
| **66** | conv3_block3_1_bn | BatchNormalization | conv3_block3_1_conv | (None, 28, 28, 128) |
| **67** | conv3_block3_1_relu | Activation | conv3_block3_1_bn | (None, 28, 28, 128) |
| **68** | conv3_block3_2_pad | ZeroPadding2D | conv3_block3_1_relu | (None, 30, 30, 128) |
| **69** | conv3_block3_2_conv | Conv2D | conv3_block3_2_pad | (None, 28, 28, 128) |
| **70** | conv3_block3_2_bn | BatchNormalization | conv3_block3_2_conv | (None, 28, 28, 128) |
| **71** | conv3_block3_2_relu | Activation | conv3_block3_2_bn | (None, 28, 28, 128) |
| **72** | conv3_block3_3_conv | Conv2D | conv3_block3_2_relu | (None, 28, 28, 512) |
| **73** | conv3_block3_out | Add | conv3_block2_out conv3_block3_3_conv | (None, 28, 28, 512) |
| **74** | conv3_block4_preact_bn | BatchNormalization | conv3_block3_out | (None, 28, 28, 512) |
| **75** | conv3_block4_preact_relu | Activation | conv3_block4_preact_bn | (None, 28, 28, 512) |
| **76** | conv3_block4_1_conv | Conv2D | conv3_block4_preact_relu | (None, 28, 28, 128) |
| **77** | conv3_block4_1_bn | BatchNormalization | conv3_block4_1_conv | (None, 28, 28, 128) |
| **78** | conv3_block4_1_relu | Activation | conv3_block4_1_bn | (None, 28, 28, 128) |
| **79** | conv3_block4_2_pad | ZeroPadding2D | conv3_block4_1_relu | (None, 30, 30, 128) |
| **80** | conv3_block4_2_conv | Conv2D | conv3_block4_2_pad | (None, 14, 14, 128) |
| **81** | conv3_block4_2_bn | BatchNormalization | conv3_block4_2_conv | (None, 14, 14, 128) |
| **82** | conv3_block4_2_relu | Activation | conv3_block4_2_bn | (None, 14, 14, 128) |
| **83** | max_pooling2d_4 | MaxPooling2D | conv3_block3_out | (None, 14, 14, 512) |
| **84** | conv3_block4_3_conv | Conv2D | conv3_block4_2_relu | (None, 14, 14, 512) |
| **85** | conv3_block4_out | Add | max_pooling2d_4 conv3_block4_3_conv | (None, 14, 14, 512) |
| **86** | conv4_block1_preact_bn | BatchNormalization | conv3_block4_out | (None, 14, 14, 512) |
| **87** | conv4_block1_preact_relu | Activation | conv4_block1_preact_bn | (None, 14, 14, 512) |
| **88** | conv4_block1_1_conv | Conv2D | conv4_block1_preact_relu | (None, 14, 14, 256) |
| **89** | conv4_block1_1_bn | BatchNormalization | conv4_block1_1_conv | (None, 14, 14, 256) |
| **90** | conv4_block1_1_relu | Activation | conv4_block1_1_bn | (None, 14, 14, 256) |
| **91** | conv4_block1_2_pad | ZeroPadding2D | conv4_block1_1_relu | (None, 16, 16, 256) |
| **92** | conv4_block1_2_conv | Conv2D | conv4_block1_2_pad | (None, 14, 14, 256) |
| **93** | conv4_block1_2_bn | BatchNormalization | conv4_block1_2_conv | (None, 14, 14, 256) |
| **94** | conv4_block1_2_relu | Activation | conv4_block1_2_bn | (None, 14, 14, 256) |
| **95** | conv4_block1_0_conv | Conv2D | conv4_block1_preact_relu | (None, 14, 14, 1024) |
| **96** | conv4_block1_3_conv | Conv2D | conv4_block1_2_relu | (None, 14, 14, 1024) |
| **97** | conv4_block1_out | Add | conv4_block1_0_conv conv4_block1_3_conv | (None, 14, 14, 1024) |
| **98** | conv4_block2_preact_bn | BatchNormalization | conv4_block1_out | (None, 14, 14, 1024) |
| **99** | conv4_block2_preact_relu | Activation | conv4_block2_preact_bn | (None, 14, 14, 1024) |
| **100** | conv4_block2_1_conv | Conv2D | conv4_block2_preact_relu | (None, 14, 14, 256) |
| **101** | conv4_block2_1_bn | BatchNormalization | conv4_block2_1_conv | (None, 14, 14, 256) |
| **102** | conv4_block2_1_relu | Activation | conv4_block2_1_bn | (None, 14, 14, 256) |
| **103** | conv4_block2_2_pad | ZeroPadding2D | conv4_block2_1_relu | (None, 16, 16, 256) |
| **104** | conv4_block2_2_conv | Conv2D | conv4_block2_2_pad | (None, 14, 14, 256) |
| **105** | conv4_block2_2_bn | BatchNormalization | conv4_block2_2_conv | (None, 14, 14, 256) |
| **106** | conv4_block2_2_relu | Activation | conv4_block2_2_bn | (None, 14, 14, 256) |
| **107** | conv4_block2_3_conv | Conv2D | conv4_block2_2_relu | (None, 14, 14, 1024) |
| **108** | conv4_block2_out | Add | conv4_block1_out conv4_block2_3_conv | (None, 14, 14, 1024) |
| **109** | conv4_block3_preact_bn | BatchNormalization | conv4_block2_out | (None, 14, 14, 1024) |
| **110** | conv4_block3_preact_relu | Activation | conv4_block3_preact_bn | (None, 14, 14, 1024) |
| **111** | conv4_block3_1_conv | Conv2D | conv4_block3_preact_relu | (None, 14, 14, 256) |
| **112** | conv4_block3_1_bn | BatchNormalization | conv4_block3_1_conv | (None, 14, 14, 256) |
| **113** | conv4_block3_1_relu | Activation | conv4_block3_1_bn | (None, 14, 14, 256) |
| **114** | conv4_block3_2_pad | ZeroPadding2D | conv4_block3_1_relu | (None, 16, 16, 256) |
| **115** | conv4_block3_2_conv | Conv2D | conv4_block3_2_pad | (None, 14, 14, 256) |
| **116** | conv4_block3_2_bn | BatchNormalization | conv4_block3_2_conv | (None, 14, 14, 256) |
| **117** | conv4_block3_2_relu | Activation | conv4_block3_2_bn | (None, 14, 14, 256) |
| **118** | conv4_block3_3_conv | Conv2D | conv4_block3_2_relu | (None, 14, 14, 1024) |
| **119** | conv4_block3_out | Add | conv4_block2_out conv4_block3_3_conv | (None, 14, 14, 1024) |
| **120** | conv4_block4_preact_bn | BatchNormalization | conv4_block3_out | (None, 14, 14, 1024) |
| **121** | conv4_block4_preact_relu | Activation | conv4_block4_preact_bn | (None, 14, 14, 1024) |
| **122** | conv4_block4_1_conv | Conv2D | conv4_block4_preact_relu | (None, 14, 14, 256) |
| **123** | conv4_block4_1_bn | BatchNormalization | conv4_block4_1_conv | (None, 14, 14, 256) |
| **124** | conv4_block4_1_relu | Activation | conv4_block4_1_bn | (None, 14, 14, 256) |
| **125** | conv4_block4_2_pad | ZeroPadding2D | conv4_block4_1_relu | (None, 16, 16, 256) |
| **126** | conv4_block4_2_conv | Conv2D | conv4_block4_2_pad | (None, 14, 14, 256) |
| **127** | conv4_block4_2_bn | BatchNormalization | conv4_block4_2_conv | (None, 14, 14, 256) |
| **128** | conv4_block4_2_relu | Activation | conv4_block4_2_bn | (None, 14, 14, 256) |

| | | | | |
|---|---|---|---|---|
| **129** | conv4_block4_3_conv | Conv2D | conv4_block4_2_relu | (None, 14, 14, 1024) |
| **130** | conv4_block4_out | Add | conv4_block3_out conv4_block4_3_conv | (None, 14, 14, 1024) |
| **131** | conv4_block5_preact_bn | BatchNormalization | conv4_block4_out | (None, 14, 14, 1024) |
| **132** | conv4_block5_preact_relu | Activation | conv4_block5_preact_bn | (None, 14, 14, 1024) |
| **133** | conv4_block5_1_conv | Conv2D | conv4_block5_preact_relu | (None, 14, 14, 256) |
| **134** | conv4_block5_1_bn | BatchNormalization | conv4_block5_1_conv | (None, 14, 14, 256) |
| **135** | conv4_block5_1_relu | Activation | conv4_block5_1_bn | (None, 14, 14, 256) |
| **136** | conv4_block5_2_pad | ZeroPadding2D | conv4_block5_1_relu | (None, 16, 16, 256) |
| **137** | conv4_block5_2_conv | Conv2D | conv4_block5_2_pad | (None, 14, 14, 256) |
| **138** | conv4_block5_2_bn | BatchNormalization | conv4_block5_2_conv | (None, 14, 14, 256) |
| **139** | conv4_block5_2_relu | Activation | conv4_block5_2_bn | (None, 14, 14, 256) |
| **140** | conv4_block5_3_conv | Conv2D | conv4_block5_2_relu | (None, 14, 14, 1024) |
| **141** | conv4_block5_out | Add | conv4_block4_out conv4_block5_3_conv | (None, 14, 14, 1024) |
| **142** | conv4_block6_preact_bn | BatchNormalization | conv4_block5_out | (None, 14, 14, 1024) |
| **143** | conv4_block6_preact_relu | Activation | conv4_block6_preact_bn | (None, 14, 14, 1024) |
| **144** | conv4_block6_1_conv | Conv2D | conv4_block6_preact_relu | (None, 14, 14, 256) |
| **145** | conv4_block6_1_bn | BatchNormalization | conv4_block6_1_conv | (None, 14, 14, 256) |
| **146** | conv4_block6_1_relu | Activation | conv4_block6_1_bn | (None, 14, 14, 256) |
| **147** | conv4_block6_2_pad | ZeroPadding2D | conv4_block6_1_relu | (None, 16, 16, 256) |
| **148** | conv4_block6_2_conv | Conv2D | conv4_block6_2_pad | (None, 7, 7, 256) |
| **149** | conv4_block6_2_bn | BatchNormalization | conv4_block6_2_conv | (None, 7, 7, 256) |
| **150** | conv4_block6_2_relu | Activation | conv4_block6_2_bn | (None, 7, 7, 256) |
| **151** | max_pooling2d_5 | MaxPooling2D | conv4_block5_out | (None, 7, 7, 1024) |
| **152** | conv4_block6_3_conv | Conv2D | conv4_block6_2_relu | (None, 7, 7, 1024) |
| **153** | conv4_block6_out | Add | max_pooling2d_5 conv4_block6_3_conv | (None, 7, 7, 1024) |
| **154** | conv5_block1_preact_bn | BatchNormalization | conv4_block6_out | (None, 7, 7, 1024) |
| **155** | conv5_block1_preact_relu | Activation | conv5_block1_preact_bn | (None, 7, 7, 1024) |
| **156** | conv5_block1_1_conv | Conv2D | conv5_block1_preact_relu | (None, 7, 7, 512) |
| **157** | conv5_block1_1_bn | BatchNormalization | conv5_block1_1_conv | (None, 7, 7, 512) |
| **158** | conv5_block1_1_relu | Activation | conv5_block1_1_bn | (None, 7, 7, 512) |
| **159** | conv5_block1_2_pad | ZeroPadding2D | conv5_block1_1_relu | (None, 9, 9, 512) |
| **160** | conv5_block1_2_conv | Conv2D | conv5_block1_2_pad | (None, 7, 7, 512) |
| **161** | conv5_block1_2_bn | BatchNormalization | conv5_block1_2_conv | (None, 7, 7, 512) |
| **162** | conv5_block1_2_relu | Activation | conv5_block1_2_bn | (None, 7, 7, 512) |
| **163** | conv5_block1_0_conv | Conv2D | conv5_block1_preact_relu | (None, 7, 7, 2048) |
| **164** | conv5_block1_3_conv | Conv2D | conv5_block1_2_relu | (None, 7, 7, 2048) |
| **165** | conv5_block1_out | Add | conv5_block1_0_conv conv5_block1_3_conv | (None, 7, 7, 2048) |
| **166** | conv5_block2_preact_bn | BatchNormalization | conv5_block1_out | (None, 7, 7, 2048) |
| **167** | conv5_block2_preact_relu | Activation | conv5_block2_preact_bn | (None, 7, 7, 2048) |
| **168** | conv5_block2_1_conv | Conv2D | conv5_block2_preact_relu | (None, 7, 7, 512) |
| **169** | conv5_block2_1_bn | BatchNormalization | conv5_block2_1_conv | (None, 7, 7, 512) |
| **170** | conv5_block2_1_relu | Activation | conv5_block2_1_bn | (None, 7, 7, 512) |
| **171** | conv5_block2_2_pad | ZeroPadding2D | conv5_block2_1_relu | (None, 9, 9, 512) |
| **172** | conv5_block2_2_conv | Conv2D | conv5_block2_2_pad | (None, 7, 7, 512) |
| **173** | conv5_block2_2_bn | BatchNormalization | conv5_block2_2_conv | (None, 7, 7, 512) |
| **174** | conv5_block2_2_relu | Activation | conv5_block2_2_bn | (None, 7, 7, 512) |
| **175** | conv5_block2_3_conv | Conv2D | conv5_block2_2_relu | (None, 7, 7, 2048) |
| **176** | conv5_block2_out | Add | conv5_block1_out conv5_block2_3_conv | (None, 7, 7, 2048) |
| **177** | conv5_block3_preact_bn | BatchNormalization | conv5_block2_out | (None, 7, 7, 2048) |
| **178** | conv5_block3_preact_relu | Activation | conv5_block3_preact_bn | (None, 7, 7, 2048) |
| **179** | conv5_block3_1_conv | Conv2D | conv5_block3_preact_relu | (None, 7, 7, 512) |
| **180** | conv5_block3_1_bn | BatchNormalization | conv5_block3_1_conv | (None, 7, 7, 512) |
| **181** | conv5_block3_1_relu | Activation | conv5_block3_1_bn | (None, 7, 7, 512) |
| **182** | conv5_block3_2_pad | ZeroPadding2D | conv5_block3_1_relu | (None, 9, 9, 512) |
| **183** | conv5_block3_2_conv | Conv2D | conv5_block3_2_pad | (None, 7, 7, 512) |
| **184** | conv5_block3_2_bn | BatchNormalization | conv5_block3_2_conv | (None, 7, 7, 512) |
| **185** | conv5_block3_2_relu | Activation | conv5_block3_2_bn | (None, 7, 7, 512) |
| **186** | conv5_block3_3_conv | Conv2D | conv5_block3_2_relu | (None, 7, 7, 2048) |
| **187** | conv5_block3_out | Add | conv5_block2_out conv5_block3_3_conv | (None, 7, 7, 2048) |
| **188** | post_bn | BatchNormalization | conv5_block3_out | (None, 7, 7, 2048) |
| **189** | post_relu | Activation | post_bn | (None, 7, 7, 2048) |
| **190** | global_average_pooling2d_1 | GlobalAveragePooling2D | post_relu | (None, 2048) |
| **191** | dropout_2 | Dropout | global_average_pooling2d_1 | (None, 2048) |
| **192** | dense_2 | Dense | dropout_2 | (None, 128) |
| **193** | dropout_3 | Dropout | dense_2 | (None, 128) |
| **194** | dense_3 | Dense | dropout_3 | (None, 2) |

### Supplementary Methods

#### Sample size and Data Characteristics

Table S2 depicts the details of the data used across the study. The current investigation employed a dataset comprising 51,274 images of rapid antigen tests, collected from 6,952 individuals. The dataset encompassed images acquired from Quidel QuickVue, BD Vertior, and Abbott BinaxNOW tests. Among these images, a total of 877 samples (1.7% of the dataset) exhibited a positive result for SARS-CoV-2. Images lacking a test card were excluded from the dataset to ensure accurate analyses (referred to as Subset 1). Subset 2 consisted of images with successfully cropped Regions of Interest (ROIs), which were used for invalid result detection and ROI classification purposes. Notably, additional ROIs from positive samples, undetected by the algorithm, were manually included in Subset 3 to retain all positive samples. Invalid test results were eliminated, resulting in the creation of Subset 3, which was subsequently utilized for training and evaluating different configurations of the ROI classification module.

#### Test Card Classification and ROI Detection

Fig. S2 presents a comprehensive workflow detailing the stages of the Test card classification module and ROI detection. Image-matching techniques were employed to classify the test cards, leveraging their ability to provide transformation information for the detected objects, thereby facilitating the subsequent step of ROI detection and cropping. Specifically, the similarity between the template image of each test card type and the inquiry image was assessed by calculating the number of correspondences, referred to as inliers, between the images. In image matching algorithms, inliers represent the reliable matched features or points that are consistent and accurate, while outliers, which are inconsistent or erroneous matches, are disregarded. These inliers are typically identified through techniques such as RANSAC (Random Sample Consensus) or other robust estimation methods.

Based on the template with the highest number of correspondences, the type of test card was determined. To ensure accurate classification, a comparison was made between the number of inliers for the best-match pair and the second-best match. If the ratio of the number of inliers for the second-best match to the best-match pair was less than 0.5, the classification was considered valid. Otherwise, the module deemed the inquiry image as an unclassifiable input. Furthermore, a mechanism was implemented to ensure uniqueness among the detected points in the destination image (inquiry image) and to guarantee that each point in the template image corresponded to a distinct point in the inquiry image. This step aimed to discard low-quality matches that may have passed the outlier removal process.

Test card classification using image matching offers several benefits over conventional object classification algorithms. There is no need for “ground truth” data, and we can avoid the time-consuming training and hyperparameter tuning process that is needed in deep learning-based object detection methods. Moreover, this module leads to a homography matrix and effortless detection of the registered ROI. Furthermore, image feature matching has been used for object detection for many years, including conventional step-by-step and recently emerging end-to-end methods [7]. Further, in this task, the precise detection of ROIs requires the extracted image features to be invariant to rotations and perspective projections and robust to changes in image properties such as brightness, contrast, and sharpness. Consequently, we assessed three image matching techniques: SIFT [1], SuperPoint + SuperGlue [2], [3], and ORB + BEBLID [4], [5] with different configurations.

After a feature matching algorithm, researchers typically use RANSAC for image matching (Fig. S2). Although RANSAC has achieved very promising results [24], [25], the model quality highly depends on the inlier-outlier threshold, unlike the setting of the upper bound threshold as in MAGSAC++ [10]. Moreover, MAGSAC++ utilizes an iterative reweighted least-squares approach, which leads to faster convergence. MAGSAC++ is not based on assumptions about the distribution of outliers and inliers, so it is likely to lead to a more robust result. Moreover, GCRANSAC (Graph-Cut RANSAC) and DEGENSAC (DEGenerate RANSAC) are two extensions of the RANSAC algorithm that address some of these limitations. GCRANSAC utilizes a graph-cut optimization technique to optimize the consensus set of inliers in each iteration, which leads to a more accurate estimation of the model parameters. DEGENSAC, on the other hand, addresses the problem of setting the RANSAC threshold manually. Instead, it uses a statistical method to estimate the threshold automatically. Both GCRANSAC and DEGENSAC have been shown to outperform RANSAC in terms of accuracy, robustness, and speed in various applications. Therefore, we assessed MAGSAC++, GCRANSAC [26], and DEGENSAC [11] for outlier removal after each feature matching algorithm (Fig. S2).

In the ROI detection module, the first step is homography estimation using the matched correspondences of the pair that has the highest number of inliers. We implemented another mechanism to verify that all the detected points in the destination image (inquiry image) are unique, to ensure that every point in the template image corresponds to a unique point in the inquiry image. This is a crucial step to drop all low-quality matches that passed the outlier removal.

SIFT

SIFT [1] is widely utilized for object recognition [8] and well-known for being scale- and rotation-invariant. To start, we used SIFT to extract and describe the features of template and inquiry image pairs. Next, the Fast Library for Approximate Nearest Neighbors (FLANN) algorithm approximately matches the feature descriptors between template image and inquiry image. Next, the first-to-second nearest neighbor ratio check (SNN) ratio test attempts to eliminate ambiguous matches such that the rest of the matches are unique. We then tested three aforementioned outlier removal methods: MAGSAC++, GCRANSAC, and DEGENSAC. The correspondences were determined to be well-matched if the number of correspondences was larger than a certain threshold ($t_{Abbott}, t_{BD}, t_{QuickVue}$) for each type of test card. For example, in Fig. S2, the inquiry image has the most number of inliers (57) in pair with the Abbott template. Since it is more than $t_{Abbott}$=10, it is valid, and since the ratio of the second-best match to the best match is less than 0.5, it is considered to be a good prediction. Therefore, matched correspondences with Abbott test cards will be sent to the next stage.

In the ROI detection module, the first step is homography estimation using matched correspondences of a pair that has the most number of inliers. we put another mechanism to verify that all the detected points in the destination image (inquiry image) are unique. In this way, we ensure that every point in the template image corresponds to a unique point in the inquiry image. This is a crucial step since in some inquiry images the SNN algorithm is not able to eliminate all low-quality matches.

To further make the model robust, after homography estimation, another mechanism is needed to verify the estimated homography matrix. In fact, in rare cases, there is a lack of robustness using MAGSAC++. Therefore, an additional homography verification step is added after the homography matrix is estimated. To remove these false correspondences, we developed an iterative algorithm to try MAGSAC++ 10 times until it can find a reasonable homography matrix. The homography matrix is a 3×3 homogeneous transformation matrix (Equation 1). In this matrix $h_{13}$ and $h_{23}$ are translation components, $h_{12}$ and $h_{21}$ are shear/rotation components, $h_{33}$ is a homogeneous scaling factor, and $h_{11}$ and $h_{22}$ are scale/rotation components which we defined as a constraint for them. Indeed, when these components are very large, the scaling has been done extremely great, therefore, the transformation result will not be good enough. Hence, if absolute values of $h_{11}$ or $h_{22}$ are less than 3, the algorithm verifies the homography matrix. Threshold 3 is concluded by experiment on the dataset. This step iterates the outlier removal algorithm till gets a better result, after ten attempts, if it is not feasible, it eliminates the inquiry image as an unclassifiable input.

$$H = \begin{bmatrix} h_{11} & h_{12} & h_{13} \\ h_{21} & h_{22} & h_{23} \\ h_{31} & h_{32} & h_{33} \end{bmatrix} \qquad \text{Equation 1}$$

SuperPoint + SuperGlue

SuperPoint is an end-to-end learning-based feature extraction algorithm that outputs the points of interest with their corresponding feature descriptors directly from a full-sized image. Unlike SIFT, SuperPoint is based on the deep neural network and therefore requires more computational resources like GPU. Moreover, SuperGlue is a learning-based matcher that utilizes graph neural networks to match local features and find correspondences. SuperPoint+SuperGlue is faster and may offer implementation advantages compared to conventional step-by-step methods such as SIFT. Further, a combination of SuperPoint and SuperGlue (SP+SG) outperformed methods such as SIFT and ORB with the nearest neighbor (NN) matcher in pose estimation and homography matrix estimation [3]. Algorithm 1 illustrates the procedure for generating the ROI using SuperPoint+SuperGlue. After obtaining destination keypoints and descriptors using SuperPoint (line 8), SuperGlue predicts matches using descriptors (line 9).

**Algorithm 1 Pseudo code of SuperPoint + SuperGlue method**

```
    Input: Template image (source image), inquiry image (Destination image), coordination of ROI in the template image, minimum number of inliers threshold
    Output: ROI of the inquiry image
1:  min_num_inliers ← 15
2:  src_im ← LoadImages(source_image_path)
3:  src_kpts, src_desc ← SuperPoint(destination_image_path)
4:  dst_im ← LoadImages(destination_image_path)
5:  all_src_mkpts ← [ ]
6:  all_dst_mkpts ← [ ]
7:  for i = 0: 4 do
8:     dst_kpts(i), dst_desc(i) ← SuperPoint(dst_im)
9:     src_mkpts(i), dst_mkpts(i) ← SuperGlue(src_kpts, src_desc, dst_kpts(i), dst_desc(i))
10:    all_src_mkpts ← Append(src_mkpts(i))
11:    all_dst_mkpts ← Append(dst_mkpts(i))
12:    dst_im ← rotateImage(dst_im, 90 degrees)
13: all_src_mkpts,  all_dst_mkpts ← OutlierRemoval(all_src_mkpts,  all_dst_mkpts)
14: If length(all_src_mkpts) > min_num_inliers then
15:    H ← HomographyEstimation(all_src_mkpts, all_dst_mkpts)
16: else
17:    Error ← " Image is unclassifiable"
18:    end procedure
19: If HomographyMatrixVerification(H) then
20:    Transformed_image ← warpPerspective(src_im, dst_im, H)
21:    ROI ← crop(transformed_im, oordination_of _ROI)
22:    Return ROI
23: else
24:    Error ← "Image is unclassifiable"
25:    end procedure
```

The SuperPoint+SuperGlue algorithm is trained with real images [9]; therefore, this method cannot handle in-plane rotations greater than 45 °. To overcome this issue, the proposed algorithm rotates the image by 90 °and performs the image matching. In this manner, the inquiry image is fed in four different orientations to the module. Then, the correspondences from all four images are assigned to the outlier removal and form the final inliers (line 13). Indeed, this modification leads to more rotation-invariant properties, which are essential for the detection of the LFA in the image. If the number of inliers is greater than the threshold (line 14), it is passed to the next stage, which is ROI detection. This stage is the same as SIFT: homography estimation (line 15), homography matrix verification (line 19), perspective transformation (line 20), and cropping (line 21). If the image cannot pass any of the aforementioned constraints, the procedure is terminated and the inquiry image is rejected as an unclassifiable input.

##### ORB + BEBLID

ORB (Oriented FAST and Rotated BRIEF), another well-known method for feature extraction and description [4], can detect keypoints in real time using FAST [10] in pyramids and further compute the descriptors using BRIEF [11]. Therefore, it can be implemented using low-power devices such as mobile phones. Moreover, it has been shown that ORB's performance is similar to that of SIFT. The Boosted Efficient Binary Local Image Descriptor (BEBLID) utilizes an ensemble learning algorithm (AdaBoost) to generate higher-quality local descriptors and is trained with imbalanced datasets to enhance the performance in asymmetric matching. It has been shown that combining ORB and BEBLID can lead to a more accurate and faster image-matching performance than just ORB.

##### Invalid Detection

To identify invalid samples, we designed a Siamese network to distinguish between the invalid and valid samples. Invalid ROIs included both unused and faulty tests which lacked the control line (Fig. S1). The proposed model is illustrated in Fig. S5. This network consists of two identical convolutional neural networks (CNN) with the same weights yet different inputs. The differences between the embeddings of two images is calculated using Euclidean distance and contrastive loss. Subsequently, the sigmoid function defines the similarity score between the images, with the final goal of predicting the similarity between the two images in each pair. If they belong to the same class, the similarity should be one; otherwise, it should be zero.

Our proposed network used a Big Transfer (m-r50x1) [6] as a feature extractor (backbone) which can learn features by leveraging a few images of invalid samples. In the prediction phase (Fig. S5), the input ROI passed through this network, and the similarity between the ROI and each invalid template was calculated separately. Consequently, if one of the similarities was more than a certain threshold, the input ROI was eliminated as an invalid sample. It is worth mentioning that, in the deployment phase, there was no need to feed the inputs as pairs. Since the template in the input pair is known, we used the feature vector (embedding) of the template along with the extracted feature vector of the inquiry ROI, and further, calculated the contrastive loss and output of the network. Therefore, in terms of computation cost, this model was equal to an equivalent solo neural network rather than two parallel networks. The summary of the proposed network architecture is elaborated in Table S7.

##### ROI Classification

The ROI Classification module aimed to classify the test result into negative and positive and provide a confidence score for its prediction. We utilized transfer learning, as many well-trained models that already have shown their capability in object classification [6]. Specifically, we finetuned ResNet 50V2 [12] and Big transfer [6] (m-r50x1) pretrained on ImageNet. Due to the challenge of handling imbalanced datasets, we conducted a series of experiments using different class weight values. Through analysis of the area under the receiver operating characteristic (ROC) curve (AUC) metric, we discovered that a 1:10 ratio of negative to positive class weight was the most effective compared to other ratios. Big Transfer utlizes Group Normalization and Weight Standardization to optimize performance which results in better performance, specifically when it comes to few-shot learning. Moreover, we tested its backbone—ResNet 50V2—with some modifications to further benchmark the performance. This modification included removing the last fully connected layers and adding two fully connected layers with two nodes as an output in order to handle the binary classification task. Table S8 and Table S9 shows the detailed layers of this architecture and its hyperparameters.

##### Model Evaluation

Test card classification and ROI detection modules were evaluated using the number of inliers and detection rate. If the detected ROI was not cropped correctly or had distortions, it was considered unclassifiable and undetected. In the deployment phase, the unclassifiable inputs were rejected based on the number of inliers. The number of inliers is a quantitative criterion that represents the confidence level of the detected ROI.

The selection of the final model was carried out utilizing a 5-fold cross-validation, with the aim of identifying the most suitable configuration for Invalid Detection and ROI classification modules. Additionally, ROC curves, sensitivity, and specificity with 95% confidence levels were used for model selection. The dataset was then partitioned into training, validation, and test sets to assess the performance of the selected model. Given the imbalanced nature of the data, threshold adjustment using the F-score and G-mean was performed during model tuning to enhance precision and generate more cautious outcomes.

The F-score, also known as the F-measure, is a performance metric that seeks to balance sensitivity and precision. To determine the optimal threshold, we calculated the F-score for each threshold value and identified the point at which this measure was maximized. Equation 2 represents the formula for this metric:

$$\mathrm{F-score} = 2 \times \frac{\mathrm{Precision\,.\ Sensitivity}}{\mathrm{Precision + Sensitivity}} \qquad \text{Equation 2}$$

G-mean is another performance metric that aims to incorporate both sensitivity and specificity. Hence, at the threshold at which the G-mean is at its peak, the sensitivity and specificity attain their maximum values. Equation 3 shows the formula for this metric.

$$\mathrm{G-mean} = \sqrt{\mathrm{Sensitivity} \times \mathrm{Specificity}} \qquad \text{Equation 3}$$

#### Experimental Setup

All experiments were conducted using desktop computers with an Intel® Xeon® CPU E5-2620 v4 @ 2.10GHz × 16 processors, two NVIDIA™ TITAN XP GPUs, and 64 Gb memory. The operating system used was Ubuntu 20.04.4 LTS, and the coding was performed using Python. TensorFlow and Keras were employed to implement deep learning models.
